# Short-term risk of falls among initiators of controlled-release tapentadol versus oxycodone: A population-based cohort study

**DOI:** 10.64898/2026.01.14.26344133

**Authors:** Ximena Camacho, Andrea L Schaffer, Jonathan Brett, Ria Hopkins, Natasa Gisev, Steven Marsh, Kristian B. Filion, Nicole Pratt, David Henry, Sallie-Anne Pearson

## Abstract

**Objective:** Given the potential severity and economic burden of falls, we compared the short-term risk of falls following initiation of sustained-release (SR) tapentadol versus controlled-release (CR) oxycodone in real-world clinical practice.

**Design:** Active comparator, new user retrospective cohort study using routinely collected health data from 2014 to 2020.

**Setting:** New South Wales, Australia.

**Participants:** People aged ≥18 years initiating publicly subsidised tapentadol (SR) or oxycodone (CR).

**Interventions:** Initiation of tapentadol (SR) or oxycodone (CR).

**Main outcome measures:** Composite measure of fall-related emergency department presentations, hospitalisations or deaths. We used propensity score matching to adjust for baseline confounding and approximated relative risks (RR) of falls at 7, 14, and 28 days after initiation using conditional logistic regression models. We calculated absolute risk differences and estimated the number of people that would need to be treated with oxycodone (CR) vs tapentadol (SR) for one additional fall to occur (NNTH). We conducted subgroup analyses restricted to people aged ≥65 and ≥80 years and by recent opioid exposure (within 90 days prior to initiation).

**Results:** We identified 103,924 tapentadol (SR) and 419,732 oxycodone (CR) initiators; after matching each cohort comprised 103,758 initiators. Most people (74%) were aged between 45-84 years, and slightly more than half were female. Within 28 days of initiation, 652 (0.6%) oxycodone (CR) initiators and 457 (0.4%) tapentadol initiators suffered a fall. Across all time points, tapentadol (SR) initiation was associated with a lower risk of falls compared with oxycodone (CR) (7 days: RR 0.57 [95% CI 0.48 to 0.69]; 14 days: 0.62 [0.54 to 0.71]; 28 days: 0.70 [0.62 to 0.79]). Absolute differences were small at all time points (approximately 1-2 fewer falls per 1,000 patients treated), corresponding to NNTH for one additional fall ranging from 739 to 529. These patterns persisted regardless of recent opioid exposure. Relative risks were similar in the older age groups while absolute differences were slightly larger (≥65 years: 2-3 fewer falls/1,000; ≥80 years: 4-8 fewer falls/1,000). The greatest absolute differences were among opioid-naïve people aged ≥80 years (6-10 fewer falls/1,000, corresponding to NNTH ranging from 173 to 97). Fall risks were attenuated among people aged ≥80 years with recent opioid exposure.

**Conclusions:** Tapentadol (SR) was associated with a lower risk of falls than oxycodone (CR) up to four weeks after initiation, although absolute differences were small. The reduction in risk may be an important consideration in older patients where the consequences of falls are most severe.

**What is already known on this topic:**

- Clinical trials suggest that sustained-release (SR) tapentadol has a lower incidence of adverse central nervous system effects compared to controlled-release (CR) oxycodone
- pioids are associated with increased risks of falls, particularly immediately following initiation

**What this study adds:**

- Initiation of tapentadol (SR) was associated with a lower short-term risk of falls resulting in ED presentation, hospitalisation, or death compared to initiation of oxycodone (CR)
- Absolute risk differences were small but reached 1% among opioid-naïve people aged 80 years and older where consequences of falls are most severe

## Introduction

Tapentadol is a µ-opioid agonist, moderate noradrenaline reuptake inhibitor (NRI) and weak serotonin reuptake inhibitor. Real-world safety of its sustained-release (SR) formulation is not yet fully understood^1, 2^. Central nervous system disorders, including dizziness and somnolence, were among the most common side effects (occurring in ≥1 in 10 people) reported in all tapentadol (SR) clinical trials^3^. Nonetheless, clinical trials of tapentadol (SR) vs controlled-release (CR) oxycodone, morphine CR, and oxycodone/naloxone combination products suggest that tapentadol (SR) has better tolerability^4^ and a lower incidence of serious adverse events than the comparator opioids^4, 5^. Both phase III and long-term safety trials have identified lower rates of dizziness and somnolence in tapentadol (SR) treatment arms vs oxycodone (CR), respectively^3, 6, 7^.

However, populations studied in clinical trials do not necessarily reflect people treated with these medicines in real-world settings. Specifically, trial participants were not permitted to use other non-study opioids. This situation does not reflect real-world clinical practice, where multiple opioids including immediate-release formulations are often used to manage pain^8^. Moreover, the trials were designed to assess efficacy in reducing pain and underpowered to assess safety outcomes. Opioids are associated with increased risks of falls compared to non-use^9, 10^; this risk is highest in the first 7 days following initiation,^11, 12^ in elderly people^10^, and persists through the first 28 days of use^11, 13^. Given the known prevalence of central nervous system side effects observed in tapentadol (SR) trials and the established link between opioid use and falls, we aimed to quantify the short-term risk of falls resulting in emergency department (ED) presentations, hospital admissions, or death, following initiation of tapentadol (SR) compared to initiation of oxycodone (CR) in real-world settings. Based on prior clinical trial evidence, we hypothesised that tapentadol (SR) would be associated with a lower incidence of falls than oxycodone (CR).

## Methods

We conducted an active comparator, new user retrospective cohort study to assess the risk of falls in individuals aged ≥18 years, in a real-world population that includes people who use other opioids alongside the opioids of interest. This study is reported in accordance with the Reporting of studies Conducted using Observational Routinely collected health Data for Pharmaco-Epidemiology (RECORD-PE) statement^14^ (Supplementary Table 1).

### Patient and Public Involvement

One of the co-investigators of this study (SM) is a consumer with lived experience being prescribed opioids (including tapentadol (SR)) to manage chronic pain. He has collaborated with us since study conception and was instrumental in helping refine the study question to focus on outcomes of tapentadol (SR) and oxycodone (CR) in a real-world context, including the inclusion of people who are prescribed multiple opioids to manage pain. He has provided input on the interpretation of findings to ensure they resonate with medicines end-users and will be involved in creating materials to disseminate study findings to patient groups and the wider community.

### Setting and data source

Australia has a universal health care system providing subsidised access to health services, including prescription medicines, and hospital care to all Australian citizens and permanent residents. This study used the Medicines Intelligence (MedIntel) Data Platform^15^. Briefly, the platform comprises longitudinal, linked population-level data for adult Australian citizens and permanent residents of New South Wales (NSW), Australia’s most populous state (8.5 million), between 2005 and 2020. The platform includes data on presentations to emergency departments (ED) and admissions to hospitals in NSW, cancer notifications, dispensing records for publicly subsidised medicines, and fact and cause of death information. More details on individual datasets are provided in Supplementary Table 2.

### Study population and study opioids

The study population comprised all adults (aged 18 years and over on the date of initiation) who initiated either tapentadol (SR) or oxycodone (CR) between 1 September 2014 and 3 December 2020.

The exposure of interest was publicly subsidised tapentadol (SR). Tapentadol prescribing (all formulations) has increased in Australia^16^, and emerging evidence suggests tapentadol is replacing oxycodone as the preferred analgesic for post-surgical pain management^17^. Although both immediate-release and sustained-release formulations are approved for use in Australia, only tapentadol (SR) is publicly subsidised. Consequently, most tapentadol sold in Australia is sustained-release, and most tapentadol (SR) sold is publicly subsidised^16, 18^. Immediate-release tapentadol is only accessible on the private market, and individual-level data on privately prescribed use were not available. Private sales of all prescribed opioids account for 27% of the overall opioid market in Australia (in oral morphine equivalent milligrams [OME])^19^.

We selected oxycodone (CR) as our comparator to align with pre-registration clinical trials of tapentadol (SR). Oxycodone (CR) was the comparator most often used in trials^4^ and most aligns with the severity and nature of pain treated with tapentadol (SR). The comparator group included both single oxycodone (CR) and combination oxycodone (CR) + naloxone products (see Supplementary Table 3 for definitions).

Tapentadol (SR) was first listed on the Pharmaceutical Benefits Scheme (PBS; Australia’s public medicines subsidy program) in June 2014. We therefore considered 1 June – 31 August 2014 as a washout period to allow for switching from private dispensing. We excluded anyone with a publicly subsidised dispensing for tapentadol (SR) during this period.

We defined initiation as a dispensing during the accrual period with no prior dispensings for either tapentadol (SR) or oxycodone (CR) in the previous 365 days. We defined the index date as the first date of initiation with either tapentadol (SR) or oxycodone (CR) during the accrual period (Supplementary Figure 1). For people who met the inclusion criteria more than once, we only included the first initiation.

We excluded people who: died or were non-residents of NSW on the day of initiation; lived in NSW for less than 1 year prior to initiation (in order to ensure sufficient data to ascertain comorbidities and history of falls); had no dispensing history in the two years prior to cohort entry; initiated both tapentadol (SR) and oxycodone (CR) on the same day; had a fall resulting in hospitalisation or ED visit in the 60 days prior to and including initiation (to minimise the risk of protopathic bias).

### Exposure and follow-up

We considered three different follow-up periods to ascertain outcomes: 7, 14, and 28 days following initiation. We chose the maximum 28-day period as it is known to be the highest risk period for falls following opioid initiation^11–13^. We followed each person from initiation until the first of: outcome of interest (fall), end of follow-up (7, 14, or 28 days), non-fall-related hospital admission (occurred in ≤0.1% of the cohort), death, or study end date (31 December 2020). We defined exposure using an intention-to-treat approach, where we assumed people were exposed to the initiated opioid (tapentadol (SR) or oxycodone (CR)) for the duration of follow-up^20^.

### Outcomes

We defined a composite measure of falls resulting in hospitalisation, ED presentation, or death (hereafter referred to as “falls”) occurring during follow-up. We ascertained falls from the hospital data using International Classification of Diseases (ICD), 10^th^ edition, Australian

Modification (ICD-10-AM); we included fall-related diagnosis codes in any position, where the fall did not occur in hospital. From ED records, we identified falls where a fall-related code was recorded as the principal diagnosis using ICD-10-AM, ICD-9, and Systematized Nomenclature of Medicine-Clinical Terms-Australian version (SNOMED-CT-AU) codes; and from death records where a fall-related ICD-10 code was recorded as either the underlying or contributing cause of death^13^ (Supplementary Table 4).

### Covariates

We included covariates that could potentially confound the association between opioids and falls, based on the literature and clinical input. We ascertained these using hospital diagnoses and dispensing claims (Supplementary Table 5). For each person, we obtained information on their demographic characteristics at the time of opioid initiation (age, sex, relative social disadvantage^21^, remoteness of residence^22^), comorbidities (cancer, history of substance abuse, any prior overdose, depression, anxiety, hypertension, atrial fibrillation or flutter, diabetes, heart failure, thyroid disease, postural hypotension, osteoporosis, stroke, renal impairment, hepatic impairment), and frailty (defined using the Hospital Frailty Risk Score^23, 24^). We also identified recent surgeries (30 days prior to initiation) and history of falls (assessed in the 2 years to 61 days prior to initiation).

We assessed exposure to non-study opioids (i.e., dispensing of any opioid other than tapentadol (SR) or oxycodone (CR)) in the 90 days prior to initiation and defined people without prior exposure as “opioid-naïve”. We also examined exposure to medicines with known risks of falls^9, 25, 26^ (falls-risk medicines; FRIDs), gabapentinoids, anticholinergic medicines^27^, and statins^28–30^ in the 30 days prior to initiation. Finally, we quantified initiation of non-study opioids on the same day as tapentadol (SR) / oxycodone (CR) initiation; initiation was defined as a dispensing for a non-study opioid with no other dispensings for that opioid in the prior year.

### Statistical analysis

We summarised the characteristics of tapentadol (SR) and oxycodone (CR) initiators using descriptive statistics and assessed group balance using standardised differences, with values >0.1 indicating meaningful imbalance^31^.

We used propensity scores to adjust for baseline confounding. We defined a propensity score model using logistic regression with the study opioid as the dependent variable and all the covariates defined above (except for initiation of non-study opioids) as independent predictors. We matched each tapentadol (SR) initiator to an oxycodone (CR) initiator (1:1) based on month and year of initiation, exposure to non-study opioids in the previous 90 days, and propensity score (using a standard caliper width of 0.2 * standard deviation of the logit of the propensity score^32^). We chose to match on propensity score (rather than weight or stratify) to carefully account for underlying trends in opioid dispensing (e.g., increasing dispensing of tapentadol over time, decreasing use of other publicly subsidised opioids, and changes to opioid prescribing policies [including codeine being moved to a prescription-only medicine])^18^ and the impact of prior opioid exposure.

#### Main analyses

We calculated separately the unadjusted relative risk (RR) under the exposure (tapentadol) and comparator (oxycodone) treatments for each outcome at 7 days, 14 days, and 28 days following initiation. We used conditional logistic regression models to approximate relative risks in the matched sample, with robust standard errors to account for the matched data. We also calculated an adjusted estimate whereby the model included a term for simultaneous initiation of study and non-study opioids. We estimated the risk differences for falls on tapentadol (SR) and oxycodone (CR) from the product of the absolute risks in the oxycodone (control) cohorts and the adjusted RR value for the relevant cohort. We estimated the number of people that would need to be treated with oxycodone (CR), rather than tapentadol (SR), for one additional fall to occur (number-needed-to-treat-to-harm; NNTH), from the inverse of the risk difference^33,34^.

#### Subgroup analyses

As risks of falls and related morbidity increase with age^13^, we created separate cohorts comprising people aged 65 years and over (65+), and people aged 80 years and older (80+) at the time of opioid initiation. We conducted our main analyses in each of these cohorts.

We also explored the risks of falls separately among people with and without exposure to non-study opioids in the 90 days prior to initiation. We conducted these analyses among our main study population and older age cohorts (65+ years, 80+ years).

#### Sensitivity analyses

We conducted several sensitivity analyses. To try to isolate the effect of the study opioids, we first restricted our study population to people who were dispensed only tapentadol (SR) or oxycodone (CR) at initiation (i.e., had no dispensings of any non-study opioids on that date). We then further restricted that cohort to people without exposure to non-study opioids in the previous 90 days.

Given that both sustained- and immediate-release opioids are often used together in practice^8^, we were interested in assessing the risk of falls in people simultaneously initiating both the study opioid and an immediate-release formulation. We restricted our study population to people who initiated both oxycodone IR (the most commonly used immediate-release opioid in Australia^19^) and study opioids on the same day.

We used a Kaplan-Meier approach to assess whether the time to falls differed between the matched tapentadol (SR) and oxycodone (CR) groups and plotted cumulative incidence curves.

We calculated the E-value^35^ to evaluate the robustness of our main findings against unmeasured confounding.

Finally, to assess the magnitude of any residual confounding we included cataract surgery as a negative control outcome. We ascertained cataract surgeries occurring during follow-up from the hospital records using Australian Classification of Health Interventions (ACHI) procedure codes (Supplementary Table 4).

All analyses were conducted with SAS v9.4 (SAS Institute, Cary NC).

## Results

We identified 103,924 tapentadol (SR) and 419,732 oxycodone (CR) initiators between 1 September 2014 and 3 December 2020; nearly three quarters were aged between 45-84 years, and slightly more than half were female. Approximately 40% of initiators of both study drugs had been dispensed non-study opioids in the prior 90 days (Table 1; Figure 1). We matched 103,758 tapentadol (SR) initiators to 103,758 oxycodone (CR) initiators on propensity scores, year and month of initiation, and exposure to non-study opioids in the prior 90 days (99% match rate).

**Table 1.**
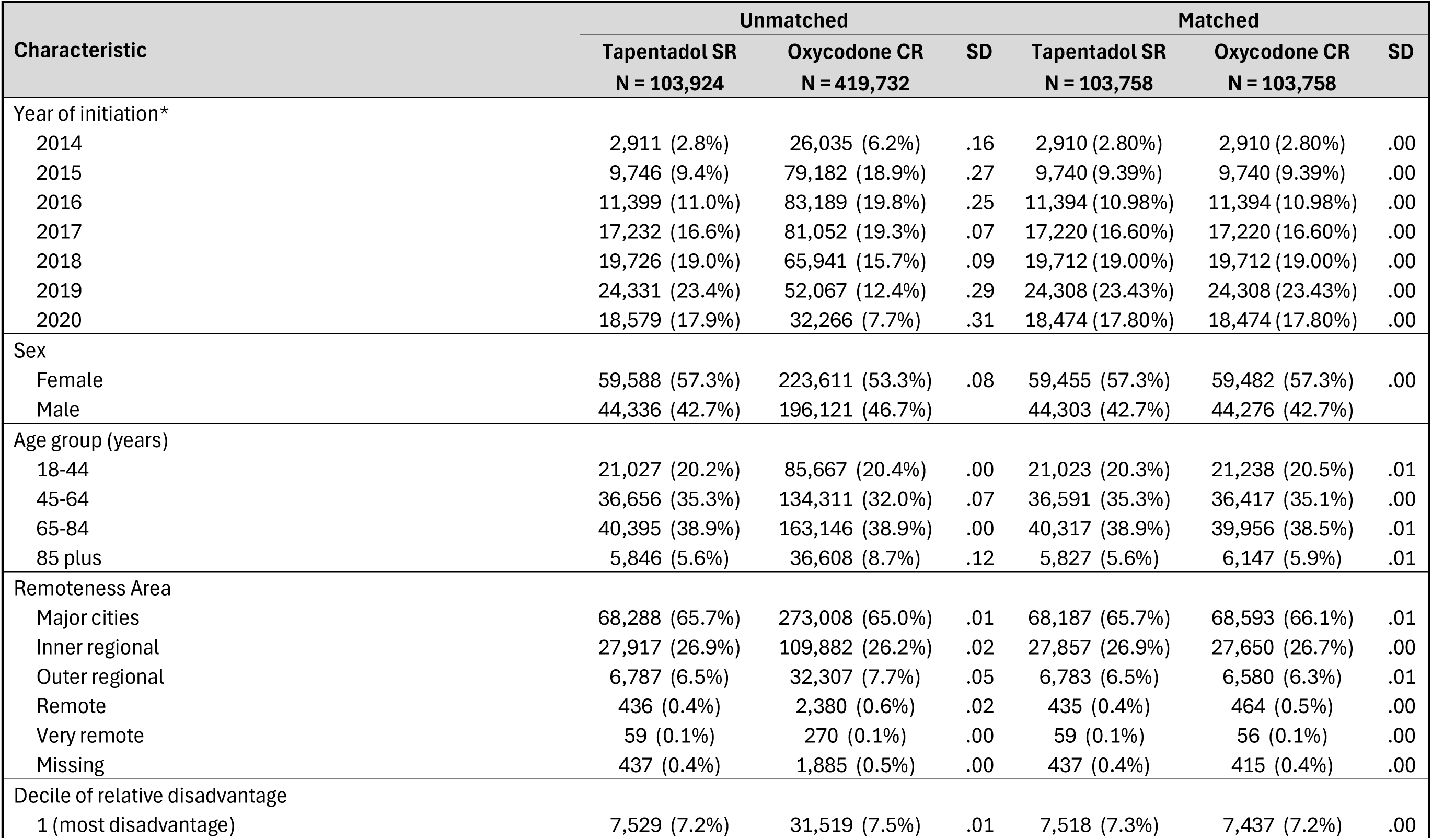
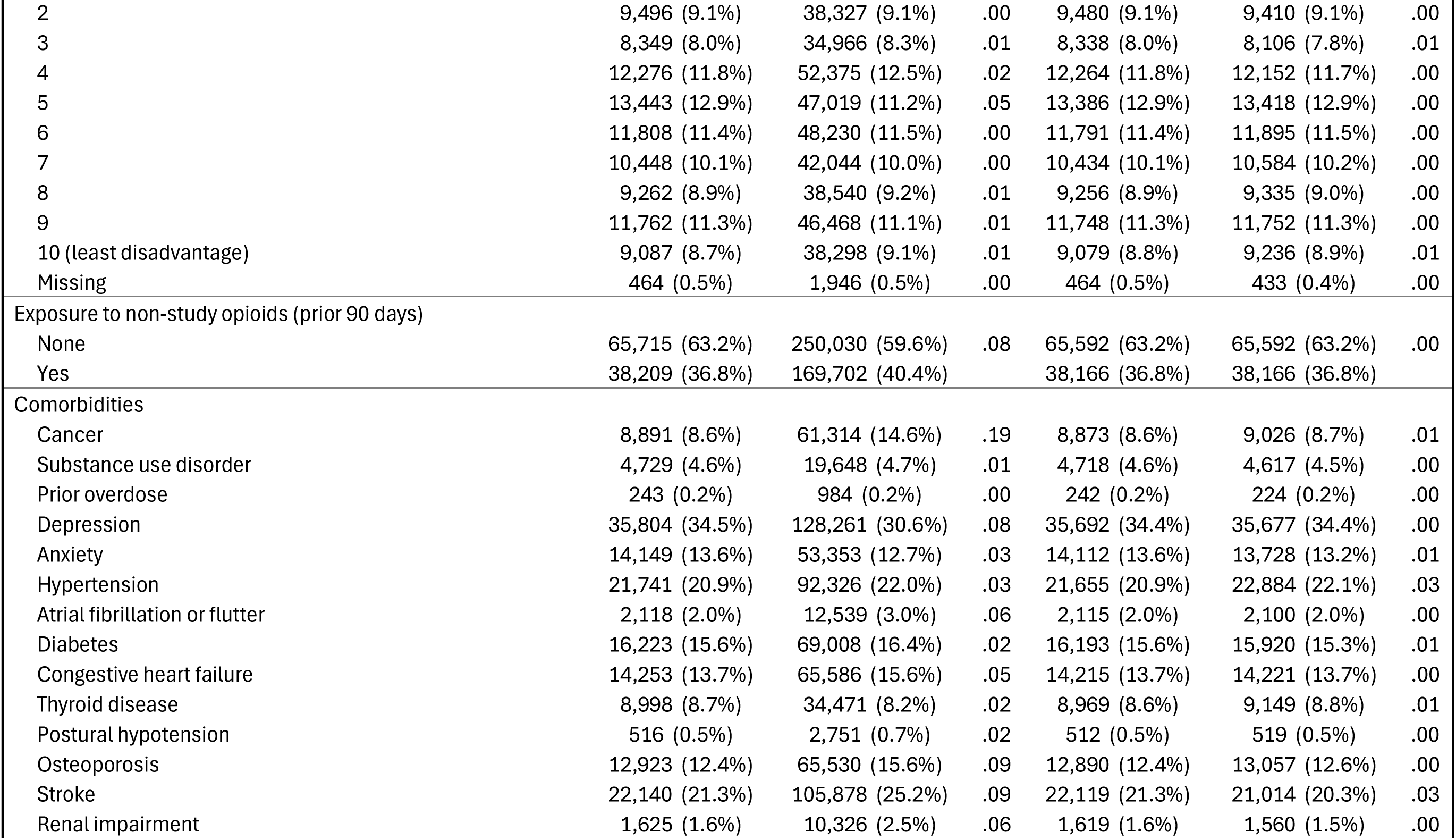
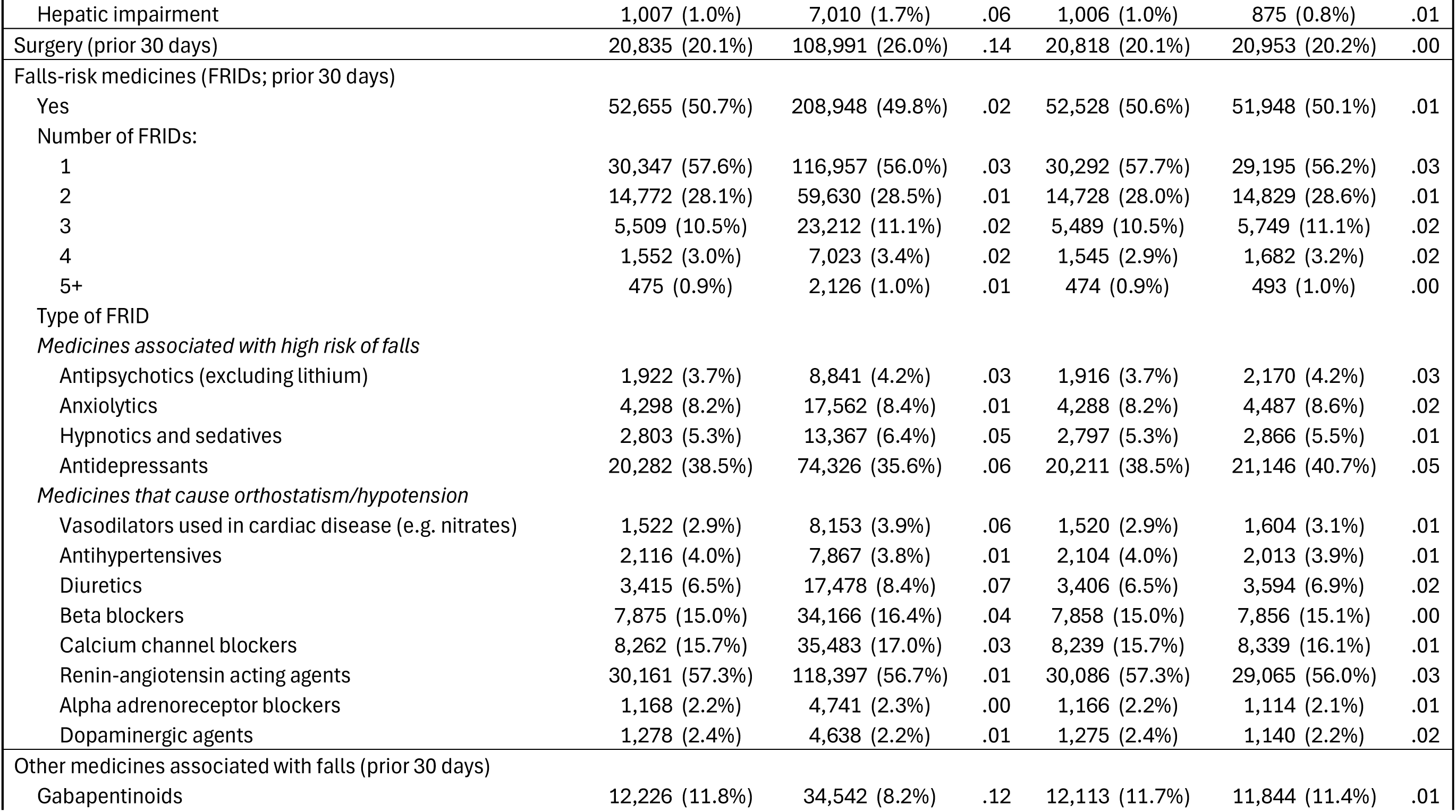
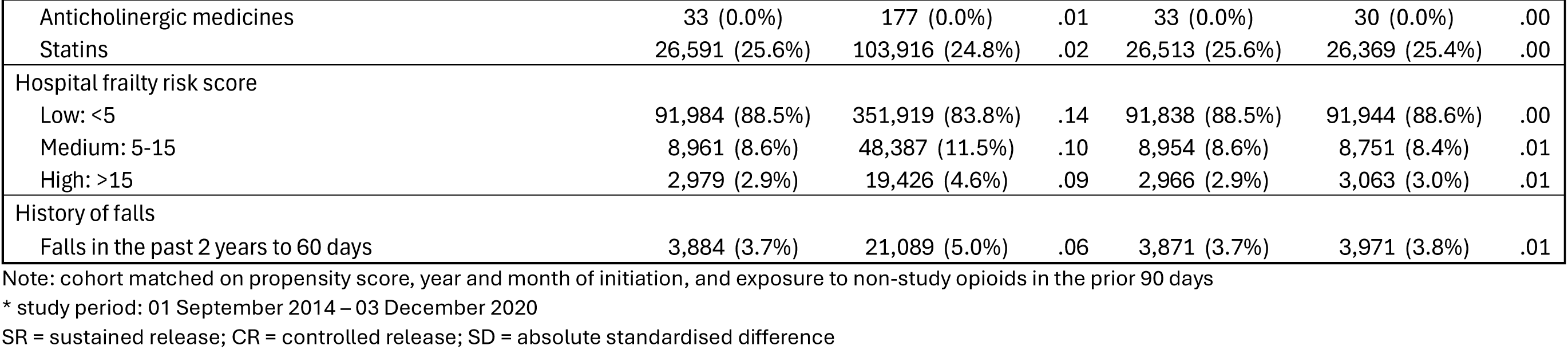
Baseline characteristics of tapentadol (SR) and oxycodone (CR) initiators, 1 September 2014 - 3 December 2020, before and after propensity score matching

**Figure 1:**
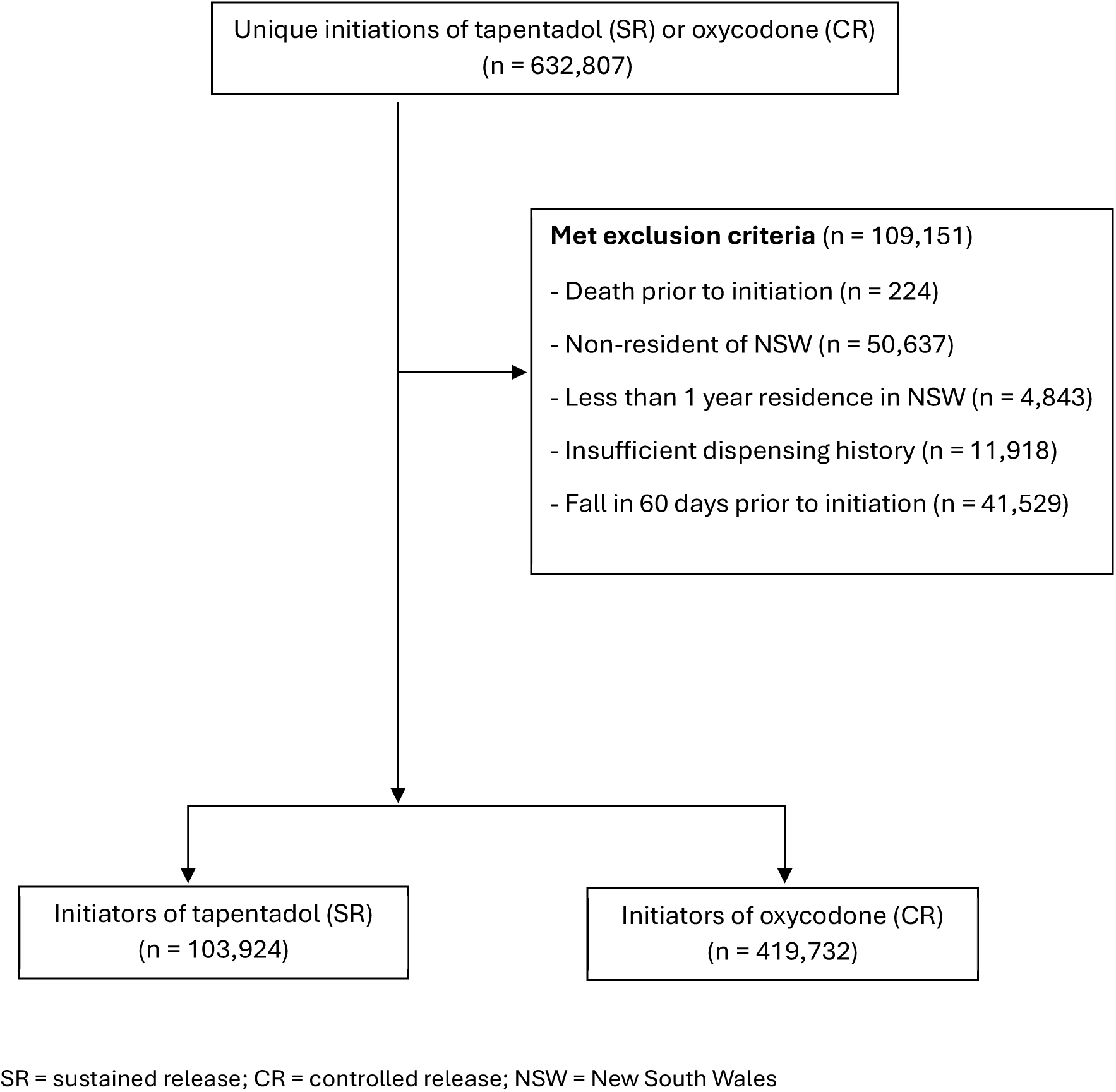
Cohort flow diagram for tapentadol (SR) and oxycodone (CR) initiators, 1 September 2014 – 3 December 2020

Covariates that were unbalanced between the two groups prior to propensity score matching were: year of initiation; age; prior cancer diagnosis; recent surgery; gabapentinoid exposure, and Hospital Frailty Risk Score. All covariates were balanced following matching.

Initiation of non-study opioids on the same day as tapentadol (SR) / oxycodone (CR) initiation differed between the two groups before and after propensity score matching (absolute standardised difference 0.36; Table 2). Immediate-release oxycodone was the most common non-study opioid dispensed, followed by codeine and tramadol.

**Table 2:** Initiation of non-study opioids at the same time as tapentadol (SR) or oxycodone (CR) initiation, for unmatched and propensity score matched cohorts

|  | Unmatched |  |  | Matched |  |  |
| --- | --- | --- | --- | --- | --- | --- |
|  | Tapentadol SR<br>N = 103,924 | Oxycodone CR<br>N = 419,732 | SD | Tapentadol SR<br>N = 103,758 | Oxycodone CR<br>N = 103,758 | SD |
| Any non-study opioid initiated | 10,657 (10.3%) | 98,981 (23.6%) | .36 | 10,646 (10.3%) | 24,523 (23.6%) | .36 |
| Type of non-study opioid initiated: |  |  |  |  |  |  |
| Codeine | 1,192 (11.2%) | 6,677 (6.8%) | .16 | 1,190 (11.2%) | 1,621 (6.6%) | .16 |
| Oxycodone | 8,470 (79.5%) | 88,239 (89.2%) | .27 | 8,462 (79.5%) | 22,058 (90.0%) | .29 |
| Fentanyl | 37 (0.4%) | 219 (0.2%) | .02 | 37 (0.4%) | 35 (0.1%) | .04 |
| Hydromorphone | 72 (0.7%) | 179 (0.2%) | .08 | 72 (0.7%) | 35 (0.1%) | .08 |
| Tramadol | 800 (7.5%) | 5,135 (5.2%) | .10 | 799 (7.5%) | 1,189 (4.9%) | .11 |
| Buprenorphine<sup>1</sup> | 238 (2.2%) | 1,085 (1.1%) | .09 | 238 (2.2%) | 260 (1.1%) | .09 |
| Morphine | 72 (0.7%) | 765 (0.8%) | .01 | 72 (0.7%) | 142 (0.6%) | .01 |
| Methadone<sup>1</sup> | 10 (0.1%) | 18 (0.0%) | .03 | 10 (0.1%) | <6 | * |

Note: cohort matched on propensity score, year and month of initiation, and exposure to non-study opioids in the prior 90 days
\* suppressed to avoid residual disclosure of small cells
<sup>1</sup> Only buprenorphine and methadone formulations with indications for pain were included in the study
SR = sustained release; CR = controlled release; SD = absolute standardised difference

### Main analyses

There were 190 (0.2%), 302 (0.3%), and 457 (0.4%) falls among tapentadol (SR) initiators at 7, 14, and 28 days following initiation, respectively. Among oxycodone (CR) initiators, there were 330 (0.3%), 487 (0.5%), and 652 (0.6%) falls at 7, 14, and 28 days post initiation, respectively.

Following propensity score matching, risks of falls were approximately 40% lower at 7 days (RR 0.57, 95% CI 0.48 to 0.69) and 14 days (RR 0.62; 95% CI 0.54 to 0.71), and were 30% lower at 28 days (RR 0.70, 95% CI 0.62 to 0.79) post initiation. Adjustment for same-day initiation of non-study opioids had minimal impact on the effect estimates (Table 3).

**Table 3:** Relative risks and risk differences for falls between propensity score matched initiators of tapentadol (SR) and oxycodone (CR), for the total study population, people aged 65+, and people aged 80+

| Outcome | Exposure | No. exposed | No. events | Matched RR<sup>1</sup><br>(95% CI) | Adjusted<sup>2</sup> RR<sup>1</sup><br>(95% CI) | Baseline risk | Risk difference<br>(95% CI) | Number needed to harm<sup>3</sup> (95% CI) |
| --- | --- | --- | --- | --- | --- | --- | --- | --- |
| Overall population |  |  |  |  |  |  |  |  |
| 7 days post-initiation | Oxycodone CR | 103,758 | 330 | 0.57 (0.48,0.69) | 0.57 (0.48,0.69) | 0.003 | -0.0014 (-0.0018,-0.0009) | 739 (556,1112) |
|  | Tapentadol SR | 103,758 | 190 |  |  |  |  |  |
| 14 days post-initiation | Oxycodone CR | 103,758 | 487 | 0.62 (0.54,0.71) | 0.61 (0.53,0.71) | 0.005 | -0.0018 (-0.0023,-0.0013) | 560 (435,770) |
|  | Tapentadol SR | 103,758 | 302 |  |  |  |  |  |
| 28 days post-initiation | Oxycodone CR | 103,758 | 652 | 0.70 (0.62,0.79) | 0.68 (0.60,0.77) | 0.006 | -0.0019 (-0.0025,-0.0013) | 529 (400,770) |
|  | Tapentadol SR | 103,758 | 457 |  |  |  |  |  |
| People aged 65 years and older |  |  |  |  |  |  |  |  |
| 7 days post-initiation | Oxycodone CR | 41,546 | 221 | 0.57 (0.46,0.71) | 0.57 (0.46,0.72) | 0.005 | -0.0023 (-0.0032,-0.0014) | 436 (313,715) |
|  | Tapentadol SR | 41,546 | 126 |  |  |  |  |  |
| 14 days post-initiation | Oxycodone CR | 41,546 | 327 | 0.60 (0.50,0.72) | 0.61 (0.51,0.73) | 0.008 | -0.0031 (-0.0042,-0.0020) | 319 (239,500) |
|  | Tapentadol SR | 41,546 | 198 |  |  |  |  |  |
| 28 days post-initiation | Oxycodone CR | 41,546 | 451 | 0.67 (0.58,0.77) | 0.67 (0.57,0.78) | 0.011 | -0.0036 (-0.0049,-0.0023) | 277 (205,435) |
|  | Tapentadol SR | 41,546 | 303 |  |  |  |  |  |
| People aged 80 years and older |  |  |  |  |  |  |  |  |
| 7 days post-initiation | Oxycodone CR | 10,783 | 122 | 0.62 (0.47,0.83) | 0.65 (0.48,0.87) | 0.011 | -0.0043 (-0.0069,-0.0017) | 233 (145,589) |
|  | Tapentadol SR | 10,783 | 76 |  |  |  |  |  |
| 14 days post-initiation | Oxycodone CR | 10,783 | 183 | 0.64 (0.51,0.81) | 0.67 (0.52,0.85) | 0.017 | -0.0061 (-0.0092,-0.0029) | 164 (109,345) |
|  | Tapentadol SR | 10,783 | 118 |  |  |  |  |  |
| 28 days post-initiation | Oxycodone CR | 10,783 | 263 | 0.68 (0.56,0.82) | 0.70 (0.57,0.85) | 0.024 | -0.0079 (-0.0115,-0.0039) | 127 (87,257) |
|  | Tapentadol SR | 10,783 | 180 |  |  |  |  |  |

<sup>1</sup> Approximated by odds ratio
<sup>2</sup> Conditional logistic regression models adjusted for simultaneous initiation of study and non-study opioids
<sup>3</sup> Number of people that would need to be treated with oxycodone (instead of tapentadol) for one additional fall to occur
SR = sustained release; CR = controlled release; RR = relative risk; CI = confidence interval

We estimated a small absolute reduction in risk of falls in the whole population treated with tapentadol (SR) compared with oxycodone (CR) across all three time periods (7 days: -0.0014, 95% CI -0.0018 to -0.0009; 14 days: -0.0018, 95% CI -0.0023 to -0.0013; 28 days: -0.0019, 95% CI -0.0025 to -0.0013). The number needed to treat with oxycodone (CR) rather than tapentadol (SR) for one additional fall to occur (NNTH) ranged from 739 (95% CI 556 to 1,112) at 7 days to 529 (95% CI 400 to 770) at 28 days after initiation (Table 3).

### Subgroup analysis: older age groups

Among initiators aged 65 years and older, crude, matched, and adjusted RR estimates were similar to those of the whole population at all time points (Table 3). Risk differences were greater in the 80+ age group (7 days: -0.0043, 95% CI -0.0069 to -0.0017; 14 days: -0.0061, 95% CI -0.0092 to -0.0029; 28 days: -0.0079, 95% CI -0.0115 to -0.0039). The estimated NNTH values in this age group ranged from 233 (95% CI 145 to 589) at 7 days to 127 (95% CI 87 to 257) at 28 days post-initiation (Table 3).

### Subgroup analysis: Risks by exposure to non-study opioids (prior 90 days)

The patterns seen in the overall population remained in our subgroup analyses. Risks of falls were lower among tapentadol (SR) initiators compared to oxycodone (CR) initiators regardless of exposure to non-study opioids in the prior 90 days (Tables 4-5).

**Table 4:** Relative risks and risk differences for falls between propensity score matched initiators of tapentadol (SR) and oxycodone (CR) with no recent exposure to non-study opioids, for the total study population, people aged 65+, and people aged 80+

| Outcome | Exposure | No. exposed | No. events | Matched RR<sup>1</sup> (95% CI) | Adjusted<sup>2</sup> RR<sup>1</sup> (95% CI) | Baseline risk | Risk difference (95% CI) | Number needed to harm<sup>3</sup> (95% CI) |
| --- | --- | --- | --- | --- | --- | --- | --- | --- |
| Overall population |  |  |  |  |  |  |  |  |
| 7 days post-initiation | Oxycodone CR | 65,592 | 202 | 0.59 (0.47,0.74) | 0.58 (0.46,0.73) | 0.003 | -0.0013 (-0.0018,-0.0007) | 787 (556,1429) |
|  | Tapentadol SR | 65,592 | 119 |  |  |  |  |  |
| 14 days post-initiation | Oxycodone CR | 65,592 | 289 | 0.62 (0.52,0.75) | 0.60 (0.50,0.73) | 0.004 | -0.0017 (-0.0023,-0.0010) | 601 (435,1000) |
|  | Tapentadol SR | 65,592 | 180 |  |  |  |  |  |
| 28 days post-initiation | Oxycodone CR | 65,592 | 392 | 0.68 (0.58,0.79) | 0.64 (0.55,0.76) | 0.006 | -0.0019 (-0.0027,-0.0011) | 522 (371,910) |
|  | Tapentadol SR | 65,592 | 267 |  |  |  |  |  |
| People aged 65 years and older |  |  |  |  |  |  |  |  |
| 7 days post-initiation | Oxycodone CR | 27,056 | 135 | 0.58 (0.43,0.76) | 0.57 (0.42,0.76) | 0.005 | -0.0021 (-0.0032,-0.0011) | 472 (313,910) |
|  | Tapentadol SR | 27,056 | 78 |  |  |  |  |  |
| 14 days post-initiation | Oxycodone CR | 27,056 | 200 | 0.57 (0.45,0.72) | 0.57 (0.45,0.73) | 0.007 | -0.0032 (-0.0045,-0.0019) | 314 (223,527) |
|  | Tapentadol SR | 27,056 | 114 |  |  |  |  |  |
| 28 days post-initiation | Oxycodone CR | 27,056 | 279 | 0.63 (0.52,0.76) | 0.61 (0.50,0.75) | 0.010 | -0.0039 (-0.0054,-0.0023) | 259 (186,435) |
|  | Tapentadol SR | 27,056 | 176 |  |  |  |  |  |
| People aged 80 years and older |  |  |  |  |  |  |  |  |
| 7 days post-initiation | Oxycodone CR | 6,812 | 86 | 0.54 (0.38,0.78) | 0.55 (0.38,0.81) | 0.013 | -0.0058 (-0.0091,-0.0024) | 173 (110,417) |
|  | Tapentadol SR | 6,812 | 47 |  |  |  |  |  |
| 14 days post-initiation | Oxycodone CR | 6,812 | 129 | 0.54 (0.40,0.72) | 0.56 (0.41,0.75) | 0.019 | -0.0087 (-0.0128,-0.0047) | 115 (79,213) |
|  | Tapentadol SR | 6,812 | 70 |  |  |  |  |  |
| 28 days post-initiation | Oxycodone CR | 6,812 | 178 | 0.60 (0.47,0.77) | 0.62 (0.48,0.80) | 0.0261 | -0.0104 (-0.0150,-0.0053) | 97 (67,189) |
|  | Tapentadol SR | 6,812 | 109 |  |  |  |  |  |

<sup>1</sup> Approximated by odds ratio
<sup>2</sup> Conditional logistic regression models adjusted for simultaneous initiation of study and non-study opioids
<sup>3</sup> Number of people that would need to be treated with oxycodone (instead of tapentadol) for one additional fall to occur
SR = sustained release; CR = controlled release; RR = relative risk; CI = confidence interval

**Table 5:**
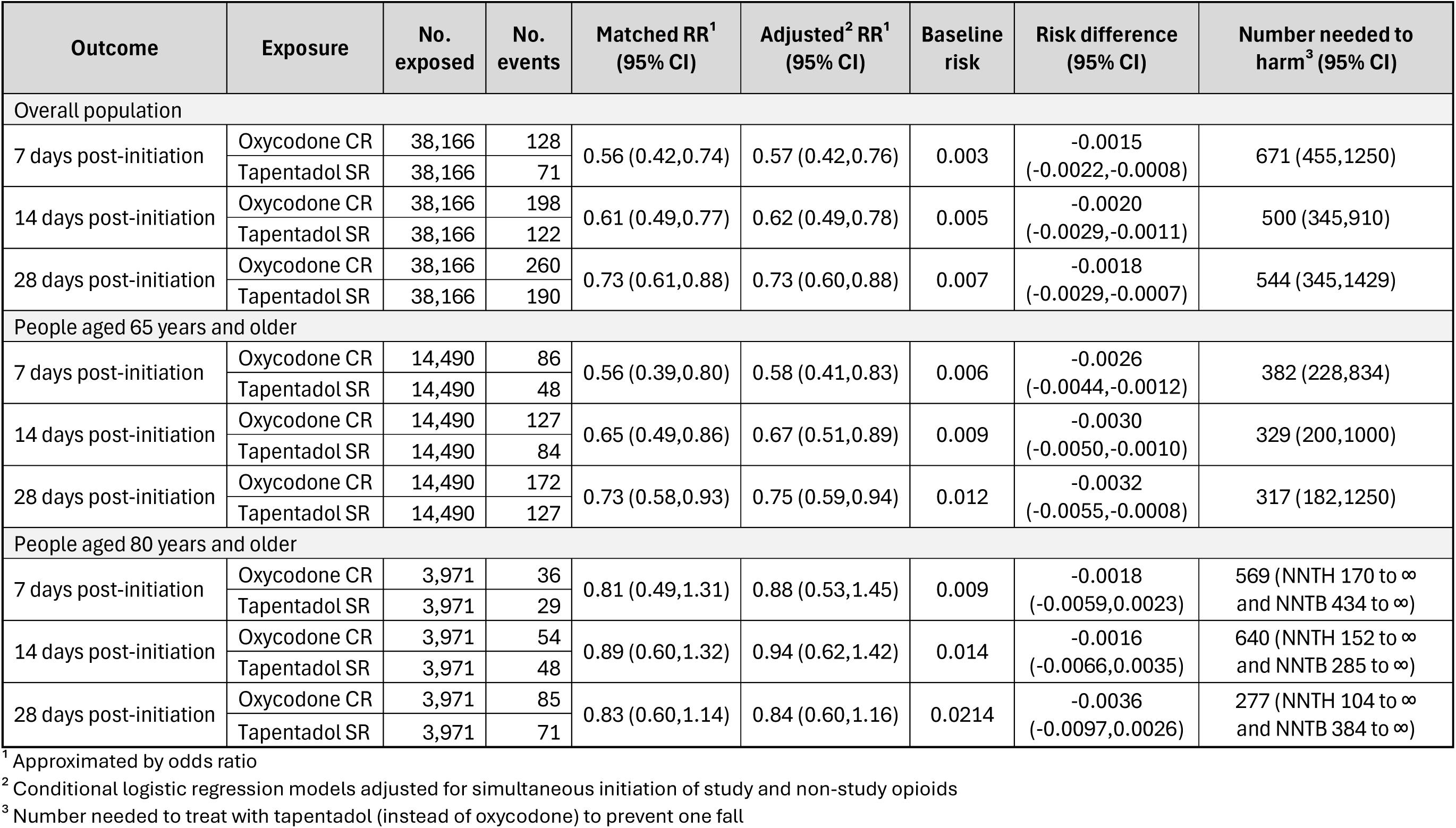
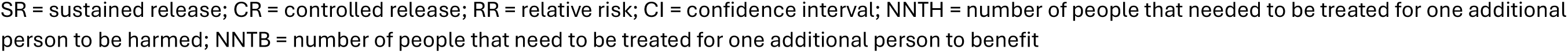
Relative risks and risk differences for falls between propensity score matched initiators of tapentadol (SR) and oxycodone (CR) with exposure to non-study opioids in the 90 days prior to initiation, for the total study population, people aged 65+, and people aged 80+

| Outcome | Exposure | No. exposed | No. events | Matched RR<sup>1</sup><br>(95% CI) | Adjusted<sup>2</sup> RR<sup>1</sup><br>(95% CI) | Baseline risk | Risk difference<br>(95% CI) | Number needed to harm<sup>3</sup> (95% CI) |
| --- | --- | --- | --- | --- | --- | --- | --- | --- |
| Overall population |  |  |  |  |  |  |  |  |
| 7 days post-initiation | Oxycodone CR | 38,166 | 128 | 0.56 (0.42,0.74) | 0.57 (0.42,0.76) | 0.003 | -0.0015<br>(-0.0022,-0.0008) | 671 (455,1250) |
|  | Tapentadol SR | 38,166 | 71 |  |  |  |  |  |
| 14 days post-initiation | Oxycodone CR | 38,166 | 198 | 0.61 (0.49,0.77) | 0.62 (0.49,0.78) | 0.005 | -0.0020<br>(-0.0029,-0.0011) | 500 (345,910) |
|  | Tapentadol SR | 38,166 | 122 |  |  |  |  |  |
| 28 days post-initiation | Oxycodone CR | 38,166 | 260 | 0.73 (0.61,0.88) | 0.73 (0.60,0.88) | 0.007 | -0.0018<br>(-0.0029,-0.0007) | 544 (345,1429) |
|  | Tapentadol SR | 38,166 | 190 |  |  |  |  |  |
| People aged 65 years and older |  |  |  |  |  |  |  |  |
| 7 days post-initiation | Oxycodone CR | 14,490 | 86 | 0.56 (0.39,0.80) | 0.58 (0.41,0.83) | 0.006 | -0.0026<br>(-0.0044,-0.0012) | 382 (228,834) |
|  | Tapentadol SR | 14,490 | 48 |  |  |  |  |  |
| 14 days post-initiation | Oxycodone CR | 14,490 | 127 | 0.65 (0.49,0.86) | 0.67 (0.51,0.89) | 0.009 | -0.0030<br>(-0.0050,-0.0010) | 329 (200,1000) |
|  | Tapentadol SR | 14,490 | 84 |  |  |  |  |  |
| 28 days post-initiation | Oxycodone CR | 14,490 | 172 | 0.73 (0.58,0.93) | 0.75 (0.59,0.94) | 0.012 | -0.0032<br>(-0.0055,-0.0008) | 317 (182,1250) |
|  | Tapentadol SR | 14,490 | 127 |  |  |  |  |  |
| People aged 80 years and older |  |  |  |  |  |  |  |  |
| 7 days post-initiation | Oxycodone CR | 3,971 | 36 | 0.81 (0.49,1.31) | 0.88 (0.53,1.45) | 0.009 | -0.0018<br>(-0.0059,0.0023) | 569 (NNTH 170 to ∞<br>and NNTB 434 to ∞) |
|  | Tapentadol SR | 3,971 | 29 |  |  |  |  |  |
| 14 days post-initiation | Oxycodone CR | 3,971 | 54 | 0.89 (0.60,1.32) | 0.94 (0.62,1.42) | 0.014 | -0.0016<br>(-0.0066,0.0035) | 640 (NNTH 152 to ∞<br>and NNTB 285 to ∞) |
|  | Tapentadol SR | 3,971 | 48 |  |  |  |  |  |
| 28 days post-initiation | Oxycodone CR | 3,971 | 85 | 0.83 (0.60,1.14) | 0.84 (0.60,1.16) | 0.0214 | -0.0036<br>(-0.0097,0.0026) | 277 (NNTH 104 to ∞<br>and NNTB 384 to ∞) |
|  | Tapentadol SR | 3,971 | 71 |  |  |  |  |  |

<sup>1</sup> Approximated by odds ratio
<sup>2</sup> Conditional logistic regression models adjusted for simultaneous initiation of study and non-study opioids
<sup>3</sup> Number needed to treat with tapentadol (instead of oxycodone) to prevent one fall
SR = sustained release; CR = controlled release; RR = relative risk; CI = confidence interval; NNTH = number of people that needed to be treated for one additional person to be harmed; NNTB = number of people that need to be treated for one additional person to benefit

The lowest risks (and largest risk differences) were among opioid-naïve adults initiating tapentadol (SR) and aged 80 years and over (Table 4), with estimated NNTH ranging from 173 (95% CI 110 to 417) to 97 (95% CI 67 to 189) at 7 and 28 days post-initiation, respectively. However, effects were attenuated among initiators aged 80+ years with exposure to opioids in the prior 90 days (Table 5).

### Sensitivity analyses

Results of our sensitivity analyses were consistent with those of our primary analysis (Supplementary Tables 10-14).

There was no notable difference in the average follow-up time between the tapentadol (SR) and oxycodone (CR) groups (Supplementary Tables 15-17; Supplementary Figures 3-5).

The E-value for falls indicated that an unmeasured confounder would have to have a RR ≥2.9 with both exposure and falls, after adjustment for all other measured confounders, to explain away the observed RR (0.57) at 7 days post-initiation. A confounder with RR ≥ 2.3 would cause the confidence interval to include 1. At 28 days post-initiation, slightly weaker confounder relationships would explain away the observed effects; an unmeasured confounder with RR ≥ 2.2 would explain away the observed RR (0.70) and one with RR ≥ 1.9 would cause the confidence interval to include 1 (Supplementary Table 18). We found similar results among our 65+ and 80+ population groups.

We did not find any differences in risks of cataract surgeries (negative control outcome) between tapentadol (SR) and oxycodone (CR) initiators at any time point or among any of our population groups (Supplementary Table 19).

## Discussion

In our population-based cohort study, we found significantly lower risks of falls resulting in ED presentations, hospitalisations, or death among people initiating tapentadol (SR) compared to those of people initiating oxycodone (CR). Tapentadol (SR) was associated with an approximately 40% lower risk of falls within the first 14 days following initiation and a 30% lower risk within 28 days of initiation. However, absolute overall risk reductions were small, with numbers-needed-to-treat with oxycodone (CR) instead of tapentadol (SR) for one additional fall to occur of 739 and 529 people at 7 and 28 days post-initiation, respectively. We found greater reductions in fall risks among older people (aged 65+ years) who were opioid-naïve. The greatest absolute reductions in fall risks were seen in opioid-naïve people aged 80+, with NNTH values for one additional fall to occur ranging from 173 to 97 people (7 and 28 days post-initiation, respectively). However, fall risks were attenuated among people aged 80+ years who had exposure to other opioids in the 90 days prior to tapentadol (SR) / oxycodone (CR) initiation.

### Comparison with other studies

Our results are consistent with findings from randomised clinical trials. Lower rates of central nervous system effects in tapentadol (SR) treatment arms have been reported in head-to-head comparisons with oxycodone (CR)^3, 6, 7^. This suggests plausibility for lower risks of falls associated with tapentadol (SR) initiation, which are likely to be mediated through these nervous system effects, particularly those affecting balance. Interestingly, Liu et al.^36^ found that tapentadol (any formulation) was associated with higher post-surgical risks of delirium compared to oxycodone (any formulation). Falls are multi-factorial and delirium is one of many potential considerations. However, delirium occurring in the immediate post-surgical period may not necessarily influence falls occurring post-discharge (the focus of our study).

Among the older age groups (65+ and 80+ years) in our study, we found that fall risks differed depending on prior exposure to non-study opioids; effect sizes were largest among people who were opioid-naïve. The risk of serious falls is highest in the first 28 days following initiation^13^; accordingly, if the true effect of tapentadol (SR) was indeed protective (in comparison to oxycodone (CR)), then presumably this would be most evident in an opioid-naïve population. Furthermore, it is reasonable to assume that a true protective effect would be more pronounced among opioid-naïve older adults where the risk of falls is highest^13^, which is consistent with our findings. We observed some attenuation of effects among older initiators (particularly people in the 80+ group) with previous exposure to non-study opioids (prior 90 days), which suggests that existing opioid tolerance may diminish any protective benefits conferred by tapentadol (SR).

### Strengths and limitations

This study leverages linked population-level data to assess outcomes of tapentadol (SR) and oxycodone (CR) in a real-world clinical setting, which includes people who are prescribed multiple opioids to manage pain.

This study had several potential limitations. First, data on dispensing of immediate-release (IR) tapentadol were not available, so people who appeared to have no opioid exposure or concurrent initiation of non-study opioids may have purchased IR tapentadol privately and be therefore misclassified. This may differentially impact our exposure groups, as people prescribed tapentadol (SR) may have been more likely to have also been prescribed tapentadol (IR). However, our findings were consistent across our main and sensitivity analyses, suggesting that the impact of misclassification was minimal. Furthermore, E-values suggested that an unmeasured confounder would require a relatively strong association with both tapentadol (SR) exposure and falls after adjustment for all measured confounders to explain away the observed relative risks. It is unlikely that tapentadol (IR) would have such a strong positive association where tapentadol (SR) appeared to have a protective effect. Nonetheless, it is critical to replicate this analysis in other data, particularly where information on tapentadol (IR) is available. Second, the data only permit observation of dispensings, but not whether medicines were actually ingested or taken as prescribed. Third, information on dose was not available in our data, and the impact of dose requires further study. Fourth, we were unable to assess duration of therapy and emulated an intention-to-treat analysis. Despite these limitations, our results were consistent across our main, subgroup and sensitivity analyses with largely constant effect sizes, and found no association between our choice of opioid and our negative control outcome (cataract surgery), lending further credibility to our findings.

### Clinical and policy implications

Falls can have devastating consequences, including mortality and life-long impacts on independence and mobility. In light of trial evidence demonstrating that tapentadol (SR) and oxycodone (CR) achieve comparable analgesic effects and improvements in functionality and quality of life^4, 37^, tapentadol (SR) may be a safer (yet equally effective) choice of pain relief, particularly among older people where the consequences of falls are most severe. Clinicians should also consider the interaction between prior opioid exposure and underlying fall risks when making opioid prescribing decisions.

Falls are the leading cause of hospitalisations and deaths due to injury, accounting for nearly a quarter of a million hospitalisations and 6,700 deaths, and costing approximately AUD $5 billion^38, 39^ in 2023/24. Although the risk reductions we found were small, with 3.1 million Australians^40^ estimated to use opioids (the majority of which is tapentadol and oxycodone^19^), and in light of the potential severity of falls outcomes, these small reductions could translate to fewer impacts on morbidity (via fewer falls) and lead to substantial health system savings.

### Conclusions

In this real-world study comparing initiators of tapentadol (SR) and oxycodone (CR), including people who use other opioids alongside the opioids of interest, we found that tapentadol (SR) was associated with a lower risk of falls resulting in ED presentation, hospitalisation, or death. Although the estimated absolute difference in risk between tapentadol (SR) and oxycodone (CR) users was small, given the widespread use of opioids this may translate to a substantial reduction in falls and associated impacts on morbidity and costs. If the effect is causal, tapentadol (SR) may be a safer and equally effective choice of opioid, particularly among older people where the consequences of falls are most severe.

## Ethics approval

This project received ethical approval from the NSW Population and Health Services Research Ethics Committee (PHSREC; approval number 2020/ETH02273) and the Australian Institute of Health and Welfare Human Research Ethics Committee (AIHW HREC; approval number EO2021/1/1233).

## Contributor information

XC, AS, JB, SM, DH, SAP conceived and designed the study. SAP acquired the data. XC performed data curation, conducted statistical analysis, and drafted the initial manuscript. All authors contributed to the interpretation of results and critically revised the manuscript. All authors reviewed and approved the final manuscript. XC and SAP are the guarantors. The corresponding author attests that all listed authors meet authorship criteria and that no others meeting the criteria have been omitted.

## Data availability statement

Direct access to the data and analytical files to other individuals or authorities is not permitted without the express permission of the approving human research ethics committees and data custodians.

## Acknowledgements

This research was completed using the Medicines Intelligence Research Program Data Platform. Data were provided by the Australian Institute of Health and Wealth (AIHW), NSW Ministry of Health and Cancer Institute NSW. Record linkage was conducted by the AIHW and Centre for Health Record Linkage (CHeReL). Secure data access was provided through the Sax Institute’s Secure Unified Research Environment (SURE).

