## Supplement for "Short-term risk of falls among initiators of controlled-release tapentadol versus oxycodone: A population-based cohort study"

### Supplementary Tables

Supplementary Table 1. The RECORD statement for pharmacoepidemiology (RECORD-PE) checklist of items, extended from the STROBE and RECORD statements, for non-interventional pharmacoepidemiological studies using routinely collected health data
Supplementary Table 2. Data sources
Supplementary Table 3. ATC and PBS item codes for identifying study opioids
Supplementary Table 4. Outcome definitions
Supplementary Table 5. Covariate definitions
Supplementary Table 6. Baseline characteristics of tapentadol (SR) and oxycodone (CR) initiators aged 65 years and older, 1 September 2014 - 3 December 2020, before and after propensity score matching
Supplementary Table 7. Baseline characteristics of tapentadol (SR) and oxycodone (CR) initiators aged 80 years and older, 1 September 2014 - 3 December 2020, before and after propensity score matching
Supplementary Table 8. Unadjusted relative risks of falls among people initiating tapentadol (SR) compared to oxycodone (CR), overall and by recent exposure to non-study opioids, for the total study population, people aged 65+, and people aged 80+
Supplementary Table 9. Baseline characteristics of tapentadol (SR) and oxycodone (CR) initiators with only study opioids dispensed on initiation date, 1 September 2014 - 3 December 2020, before and after propensity score matching
Supplementary Table 10. Relative risks of falls and cataract surgeries among tapentadol (SR) and oxycodone (CR) initiators with only study opioids dispensed on initiation date
Supplementary Table 11. Baseline characteristics of tapentadol (SR) and oxycodone (CR) initiators with only study opioids dispensed on initiation date and no exposure to non-study opioids in the previous 90 days, before and after propensity score matching
Supplementary Table 12. Relative risks of falls and cataract surgeries among tapentadol (SR) and oxycodone (CR) initiators with only study opioids dispensed on initiation date and no exposure to non-study opioids in the prior 90 days
Supplementary Table 13. Baseline characteristics of tapentadol (SR) and oxycodone (CR) initiators between 1 September 2014 - 3 December 2020 who concurrently initiated oxycodone (IR), before and after propensity score matching
Supplementary Table 14. Relative risks of falls and cataract surgeries among tapentadol (SR) and oxycodone (CR) initiators who concurrently initiated oxycodone (IR)
Supplementary Table 15. Average follow-up time (days), by exposure
Supplementary Table 16. Average follow-up time (days), by exposure, among initiators with no recent opioid exposure
Supplementary Table 17. Average follow-up time (days), by exposure, among initiators with recent exposure to opioids
Supplementary Table 18. E-values for relative risks of falls

Supplementary Table 19. Relative risks of cataract surgeries (negative control outcome) for the total study population, people aged 65+, and people aged 80+

**<u>Supplementary Figures</u>**

Supplementary Figure 1. Study design
Supplementary Figure 2. Propensity score distributions for tapentadol (SR) and oxycodone (CR) initiators
Supplementary Figure 3. Cumulative incidence curves: falls following initiation of tapentadol (SR) or oxycodone (CR)
Supplementary Figure 4. Cumulative incidence curves: falls following initiation of tapentadol (SR) or oxycodone (CR) among initiators with no recent opioid exposure
Supplementary Figure 5. Cumulative incidence curves: falls following initiation of tapentadol (SR) or oxycodone (CR) among initiators with recent exposure to opioids

**Supplementary Table 1. The RECORD statement for pharmacoepidemiology (RECORD-PE) checklist of items, extended from the STROBE and RECORD statements, for non-interventional pharmacoepidemiological studies using routinely collected health data**

| Item No | STROBE items | RECORD items | RECORD-PE[1] items | Page No |
|---|---|---|---|---|
| **Title and abstract** | | | | |
| 1 | (a) Indicate the study's design with a commonly used term in the title or the abstract.<br>(b) Provide in the abstract an informative and balanced summary of what was done and what was found. | 1.1: The type of data used should be specified in the title or abstract. When possible, the name of the databases used should be included.<br>1.2: If applicable, the geographical region and timeframe within which the study took place should be reported in the title or abstract.<br>1.3: If linkage between databases was conducted for the study, this should be clearly stated in the title or abstract. | — | 1<br>3-4 |
| **Introduction** | | | | |
| Background rationale | | | | |
| 2 | Explain the scientific background and rationale for the investigation being reported. | — | — | 6 |
| Objectives | | | | |
| 3 | State specific objectives, including any prespecified hypotheses. | — | — | 6 |
| **Methods** | | | | |
| Study design | | | | |
| 4 | Present key elements of study design early in the paper. | — | 4.a: Include details of the specific study design (and its features) and report the use of multiple designs if used.<br>4.b: The use of a diagram(s) is recommended to illustrate key aspects of the study design(s), including exposure, washout, lag and observation periods, and covariate definitions as relevant. | 7<br>Supp Fig 1 |
| Setting | | | | |
| 5 | Describe the setting, locations, and relevant dates, including periods of | — | — | 7-9 |

| | | | | |
|---|---|---|---|---|
| | recruitment, exposure, follow-up, and data collection. | | | |
| Participants | | | | |
| 6 | a) Cohort study—give the eligibility criteria, and the sources and methods of selection of participants. Describe methods of follow-up. Case-control study—give the eligibility criteria, and the sources and methods of case ascertainment and control selection. Give the rationale for the choice of cases and controls. Cross sectional study—give the eligibility criteria, and the sources and methods of selection of participants.<br>(b) Cohort study—for matched studies, give matching criteria and number of exposed and unexposed. Case-control study—for matched studies, give matching criteria and the number of controls per case. | 6.1: The methods of study population selection (such as codes or algorithms used to identify participants) should be listed in detail. If this is not possible, an explanation should be provided.<br>6.2: Any validation studies of the codes or algorithms used to select the population should be referenced. If validation was conducted for this study and not published elsewhere, detailed methods and results should be provided.<br>6.3: If the study involved linkage of databases, consider use of a flow diagram or other graphical display to demonstrate the data linkage process, including the number of individuals with linked data at each stage. | 6.1.a: Describe the study entry criteria and the order in which these criteria were applied to identify the study population. Specify whether only users with a specific indication were included and whether patients were allowed to enter the study population once or if multiple entries were permitted. See explanatory document for guidance related to matched designs. | 8-9, 11 Supp Table 3 |
| Variables | | | | |
| 7 | Clearly define all outcomes, exposures, predictors, potential confounders, and effect modifiers. Give diagnostic criteria, if applicable. | 7.1: A complete list of codes and algorithms used to classify exposures, outcomes, confounders, and effect modifiers should be provided. If these cannot be reported, an explanation should be provided. | 7.1.a: Describe how the drug exposure definition was developed.<br>7.1.b: Specify the data sources from which drug exposure information for individuals was obtained.<br>7.1.c: Describe the time window(s) during which an individual is considered exposed to the drug(s). The rationale for selecting a particular time window should be provided. The extent of potential left truncation or left censoring should be specified.<br>7.1.d: Justify how events are attributed to current, prior, ever, or cumulative drug exposure. | 8-10 Supp Tables 3-5 |

| | | | | |
|---|---|---|---|---|
| | | | 7.1.e: When examining drug dose and risk attribution, describe how current, historical or time on therapy are considered.<br>7.1.f: Use of any comparator groups should be outlined and justified.<br>7.1.g: Outline the approach used to handle individuals with more than one relevant drug exposure during the study period. | |
| Data sources/measurement | | | | |
| 8 | For each variable of interest, give sources of data and details of methods of assessment (measurement). Describe comparability of assessment methods if there is more than one group. | — | 8.a: Describe the healthcare system and mechanisms for generating the drug exposure records. Specify the care setting in which the drug(s) of interest was prescribed. | 7-10 Supp Tables 3-5 |
| Bias | | | | |
| 9 | Describe any efforts to address potential sources of bias. | — | — | 11-13 |
| Study size | | | | |
| 10 | Explain how the study size was arrived at. | — | — | 7-11 |
| Quantitative variables | | | | |
| 11 | Explain how quantitative variables were handled in the analyses. If applicable, describe which groupings were chosen, and why. | — | — | 11 Supp Table 5 |
| Statistical methods | | | | |
| 12 | (a) Describe all statistical methods, including those used to control for confounding.<br>(b) Describe any methods used to examine subgroups and interactions.<br>(c) Explain how missing data were addressed. | — | 12.1.a: Describe the methods used to evaluate whether the assumptions have been met.<br>12.1.b: Describe and justify the use of multiple designs, design features, or analytical approaches. | 11-13 |

| | | | | |
|---|---|---|---|---|
| | (d) Cohort study—if applicable, explain how loss to follow-up was addressed. Case-control study—if applicable, explain how matching of cases and controls was addressed. Cross sectional study—if applicable, describe analytical methods taking account of sampling strategy.<br>(e) Describe any sensitivity analyses. | | | |
| Data access and cleaning methods | | | | |
| 12 | — | 12.1: Authors should describe the extent to which the investigators had access to the database population used to create the study population.<br>12.2: Authors should provide information on the data cleaning methods used in the study. | — | 9 |
| Linkage | | | | |
| 12 | — | 12.3: State whether the study included person level, institutional level, or other data linkage across two or more databases. The methods of linkage and methods of linkage quality evaluation should be provided. | — | 7 |
| **Results** | | | | |
| Participants | | | | |
| 13 | (a) Report the numbers of individuals at each stage of the study (eg, numbers potentially eligible, examined for eligibility, confirmed eligible, included in the study, completing follow-up, and analysed).<br>(b) Give reasons for non-participation at each stage.<br>(c) Consider use of a flow diagram. | 13.1: Describe in detail the selection of the individuals included in the study (that is, study population selection) including filtering based on data quality, data availability, and linkage. The selection of included individuals can be described in the text or by means of the study flow diagram. | — | 13<br>Fig 1 |
| Descriptive data | | | | |
| 14 | (a) Give characteristics of study participants (eg, demographic, clinical, | — | — | 13 |

| | | | | |
|---|---|---|---|---|
| | social) and information on exposures and potential confounders.<br>(b) Indicate the number of participants with missing data for each variable of interest.<br>(c) Cohort study—summarise follow-up time (eg, average and total amount). | | | Tables 1-2 Supp Table 15 |
| Outcome data | | | | |
| 15 | Cohort study—report numbers of outcome events or summary measures over time. Case-control study—report numbers in each exposure category, or summary measures of exposure. Cross sectional study—report numbers of outcome events or summary measures. | — | — | 13 Table 3 |
| Main results | | | | |
| 16 | (a) Give unadjusted estimates and, if applicable, confounder adjusted estimates and their precision (eg, 95% confidence intervals). Make clear which confounders were adjusted for and why they were included.<br>(b) Report category boundaries when continuous variables are categorised.<br>(c) If relevant, consider translating estimates of relative risk into absolute risk for a meaningful time period. | — | — | 14 Table 3 Supp Table 8 |
| Other analyses | | | | |
| 17 | Report other analyses done—eg, analyses of subgroups and interactions, and sensitivity analyses. | — | — | 14-15 Tables 3-5 Supp Tables |

| | | | | |
|---|---|---|---|---|
| | | | | 10-14, 18, 19 |
| **Discussion** | | | | |
| Key results | | | | |
| 18 | Summarise key results with reference to study objectives | — | — | 15-16 |
| Limitations | | | | |
| 19 | Discuss limitations of the study, taking into account sources of potential bias or imprecision. Discuss both direction and magnitude of any potential bias. | 19.1: Discuss the implications of using data that were not created or collected to answer the specific research question(s). Include discussion of misclassification bias, unmeasured confounding, missing data, and changing eligibility over time, as they pertain to the study being reported. | 19.1.a: Describe the degree to which the chosen database(s) adequately captures the drug exposure(s) of interest. | 17-18 |
| Interpretation | | | | |
| 20 | Give a cautious overall interpretation of results considering objectives, limitations, multiplicity of analyses, results from similar studies, and other relevant evidence. | — | 20.a: Discuss the potential for confounding by indication, contraindication or disease severity or selection bias (healthy adherer/sick stopper) as alternative explanations for the study findings when relevant. | 16-18 |
| Generalisability | | | | |
| 21 | Discuss the generalisability (external validity) of the study results. | — | — | 17-18 |
| **Other information** | | | | |
| Funding | | | | |
| 22 | Give the source of funding and the role of the funders for the present study and, if applicable, for the original study on which the present article is based. | — | — | 2 |
| Accessibility of protocol, raw data, and programming code | | | | |
| 22 | — | 22.1: Authors should provide information on how to access any supplemental information such as the study protocol, raw data, or programming code. | — | 19 |

RECORD=reporting of studies conducted using observational routinely collected data; RECORD-PE=RECORD for pharmacoepidemiological research; STROBE=strengthening the reporting of observational studies in epidemiology.

**Supplementary Table 2. Data sources**

This study leveraged the Medicines Intelligence Data Platform, held by the Medicines Intelligence Research Program at the School of Population Health, UNSW Sydney and described elsewhere[2]. The Medicines Intelligence Data Platform comprises longitudinal, linked population-level data for adults residing in NSW between 2005 and 2020 who were eligible for Medicare (i.e., registered in the Medicare Consumer Directory).

Data dictionaries for NSW data are available from the Centre for Health Record Linkage (CHeReL)[3]. The CHeReL supports secure access to linked health data about people in New South Wales (NSW) and the Australian Capital Territory (ACT) for researchers, planners, and policy makers. The CHeReL is a node of the Population Health Research Network, which is an Australian Government initiative being conducted as part of the National Collaborative Research Infrastructure Strategy. The core functions of the CHeReL are funded by the NSW Ministry of Health.

Information on Commonwealth data collections are available from the Australian Institute for Health and Welfare (AIHW)[4]. The AIHW is a statutory Australian Government agency providing information and statistics to support policy and service delivery decisions.

| Data source | Description |
|---|---|
| NSW data collections | |
| NSW Admitted Patient Data Collection (APDC) | A statutory data collection containing records of all hospital separations (including public hospitals, public psychiatric hospitals, public multi-purpose services, private hospitals, and private day procedure centres) in NSW. Data on diagnoses are coded according to the International Classification of Diseases, Version 10, Australian Modification (ICD-10-AM) and procedures coded according to the Australian Classification of Health Interventions (ACHI). |
| NSW Emergency Department Data Collection (EDDC) | A data collection containing all visits to participating emergency departments in NSW. Data on diagnoses are coded according to the International Classification of Diseases, Version 9/10 or SNOMED-CT-AU. |
| NSW Cancer Registry (NSWCR) | A statutory data collection containing all notifications of primary malignant neoplasms from 1 Jan 1972. |
| Commonwealth data collections | |
| Medicare Consumer Directory (MCD) | Listing of individuals registered with Medicare (Australian citizens, permanent residents, and foreigners with reciprocal health care arrangements). |
| Pharmaceutical Benefits Scheme (PBS)[5] | Claims for dispensed prescription medicines covered under the Pharmaceutical Benefits Scheme. All Medicare-eligible residents are eligible to receive medicines subsidised by the PBS. The PBS covers dispensings from community pharmacies and private hospitals; information on medicines provided in acute care, emergency departments and by private prescriptions is not available. |
| National Death Index (NDI)[6] | Listing of all deaths (with cause of death information) registered in Australia since 1980. |

NSW = New South Wales; SNOMED-CT-AU: Systematized Nomenclature of Medicine-Clinical Terms-Australian version.

**Supplementary Table 3. ATC and PBS item codes for identifying study opioids**

All exposures are ascertained from the Pharmaceutical Benefits Scheme (PBS) dispensing records.

| Exposure | ATC code | PBS item codes |
|---|---|---|
| Tapentadol (SR) | N02AX06 | 10096J, 10094G, 10100N, 10091D, 10092E |
| Oxycodone (CR) | N02AA05 | 08385H, 08386J, 08387K, 08388L, 08681X, 09399Q, 09400R, 12510K, 12518W, 12525F, 12527H, 12538X, 12545G, 05015Y, 05016B, 05227D, 05247E, 05248F, 05249G, 05250H, 06058W, 06059X, 06060Y, 06061B, 06079Y, 07215R, 07216T |
| | N02AA55 | 08000C, 08934F, 08935G, 08936H, 10757E, 10758F, 10776E, 11102H, 11111T, 12471J, 12475N, 12486E, 12498T, 12511L, 12522C, 12523D, 12532N, 12540B |

ATC: Anatomical Therapeutic Chemical classification; PBS: Pharmaceutical Benefits Scheme.

**Supplementary Table 4. Outcome definitions**

Falls

Falls were defined as any hospitalisation, emergency department presentation, or death related to a fall, occurring during the follow-up period[7-9].

*Hospital data:* any ICD-10-AM code in any diagnosis field (principal or other) where the fall was not classified as an in-hospital event (i.e., condition onset flag ^= 1 [condition with onset during the episode of care]).

*Emergency department data:* any ICD-9 or SNOMED-CT-AU code in any diagnosis field.

*Death data:* any ICD-10 code in any cause of death field (underlying or other cause of death).

Cataract surgery

Cataract surgeries were defined as any hospitalisation related to cataract surgery occurring during the follow-up period. A procedure block or code in any position was sufficient to ascertain the event of interest[10].

| Outcome | Data source | Codes |
| --- | --- | --- |
| Falls resulting in hospitalisation, ED presentation, or death | ICD-10-AM code in EDDC or APDC | W00, W01, W02, W03, W04, W05, W06, W07, W08, W09, W10, W11, W12, W13, W14, W15, W16, W17, W18, W19 |
| | ICD-10 code in death records | W00, W01, W02, W03, W04, W05, W06, W07, W08, W09, W10, W11, W12, W13, W14, W15, W16, W17, W18, W19 |
| | ICD-9 code in EDDC | E880, E881, E882, E883, E884, E885, E886, E887, E888 |

| Outcome | Data source | Codes |
|---|---|---|
| | SNOMED-CT-AU code in EDDC | 298343000, 249994007, 298345007, 61683000, 161898004, 252318014, 298344006, 439017019, 408561005, 720141000168100, 249997000, 298346008, 439020010, 298347004, 439021014, 279992002, 417481019, 404911003, 2156786012, 249995008, 1144480005, 404912005, 2156787015, 363801007, 363802000, 298348009, 427206005, 398117008, 18890001000004100, 32121000119107, 242177005, 218250005, 1912002, 4301012, 217082002, 329287012, 329286015, 715157007, 715158002, 68062003, 13935005, 242399008, 242400001, 242401002, 242398000, 362715011, 57741007, 217173005, 217147000, 217106003, 242413007, 242417008, 20902002, 83468000, 138437018, 217142006, 242415000, 713397003, 242414001, 56307009, 242419006, 17886000, 217148005, 242126003, 429482004, 86591008, 217097004, 84026000, 242408008, 14047009, 217103006, 242409000, 242410005, 217107007, 217108002, 217112008, 242411009, 4521008, 242412002, 2617007, 217110000, 41411008, 72738009, 78331008, 13711007, 13278006, 217150002, 82947003, 217109005, 90639005, 40104005, 66810014, 74541001, 90619006, 44188002, 439570003, 216107001, 216108006, 216116002, 216111007, 216117006, 216113005, 216110008, 216115003, 216119009, 216123001, 216129002, 216125008, 216122006, 216127000, 216128005, 216154002, 242228005, 242227000, 216157009, 216163000, 216159007, 216156000, 216161003, 216162005, 216131006, 216135002, 216141009, 216137005, 216134003, 216140005, 242423003, 217130001, 68274007, 217124001, 217126004, 242420000, 242421001, 242426006, 242428007, 242427002, 217132009, 217133004, 217134005, 242425005, 242424009, 217135006, 242422008, 217123007, 39109007, 78331008, 242182003, 242183008, 242184002, 215644005, 215651001, 215646007, 215645006, 215650000, 215634008, 215641002, 215636005, 215635009, 215640001, 242185001, 75354000, 217083007, 217084001, 217086004, 217088003, 217090002, 414188008, 242109009, 214436006, 214437002, 214439004, 214441003, 214438007, 214443000, 214444006, 214442005, 414190009, 217092005, 217094006, 2534632013, 217093000, 242120009, 242122001, 242121008, 242123006, 33036003, 274919008, 288296009, 242402009, 217161005, 217162003, 274918000, 242397005, 217155007, 242391006, 242390007, 242387001, 242388006, 242389003, 242392004, 217157004, 217156008, 242394003, 242393009, 242395002, 242396001, 217158009, 269699007, 242404005, 242405006, 242406007, 242407003, 217154006, 215633002, 418111002, 48015001, 60594001, 242125004, 225054009, 78361000, 54670004, 82612009, 75941004, 213911003, 213925002, 242078003, 242079006, 213927005, 213926001, 213912005, 213914006, 213913000, 213917004, 213919001, 213922004, 213920007, 213918009, 429636000, 427849003, 429621003, 67223001, 56962005, 429012000, 215295004, 215298002, 215296003, 215297007, 242111000, 242112007, 214447004, 214448009, 214450001, 214452009, 214449001, 214455006, 214453004, 214538005, 214993007, 215227000, 18890001000004102, 217924002, 414189000 |

| Outcome | Data source | Codes |
|---|---|---|
| Cataract surgery | ACHI block in APDC | Data years 2015/16, 2016/17: 195, 196, 197, 198, 199, 200, 201<br>Data years 2017/18: 200<br>Data years 2018/19: 200<br>Data years 2019/20, 2020/21: 200 |
| | ACHI procedure code in APDC | Data years 2014/15: 42698-00 to 42698-05, 42702-00 to 42702-11, 42716-00, 42719-00, 42719-02, 42722-00, 42731-00, 42731-01, 42734-00, 42788-00<br>Data years 2017/18: 42734-01, 90077-00<br>Data years 2018/19: 42737-01, 42734-01<br>Data years 2019/20, 2020/21: 42737-01, 42734-01 |

APDC: Admitted Patient Data Collection; ACHI: Australian Classification of Health Interventions; EDDC: Emergency Department Data Collection; ICD-9: International Statistical Classification of Diseases and Related Health Problems, 9th Revision; ICD-10: International Classification of Diseases, 10th Revision; ICD-10-AM: International Classification of Diseases, 10th Revision, Australian Modification; SNOMED-CT-AU: Systematized Nomenclature of Medicine-Clinical Terms-Australian version.

**Supplementary Table 5. Covariate definitions**

***Relative disadvantage:*** We used a relative area-based socio-economic index based on Census data (Index of Relative Socio-Economic Disadvantage; IRSD) that summarises a range of information about economic and social conditions. The score is based on a combination of factors such as income, education, employment, housing, and family structure. Scores are calculated for each Statistical Area; deciles are then created by ordering the areas by disadvantage and dividing into ten equally sized groups, with decile 1 containing the most disadvantaged areas and decile 10 the most advantaged[11]. For each initiator, we used the Socio-Economic Index for Australia (SEIFA) correspondence file from the Australian Bureau of Statistics (ABS) to map the postal code at initiation to the corresponding IRSD decile (2016 SEIFA index).

***Remoteness:*** We defined remoteness using the Australian Statistical Geographic Standard Remoteness Structure[12]. This structure divides Australia into five classes of remoteness characterised by relative geographic access to services, measured using the Accessibility/Remoteness Index of Australia Plus (ARIA+)[13]. For each initiator, we used an ABS correspondence file to map the postal code at the time of initiation to the corresponding remoteness area (2016 Remoteness Area structure).

***Hospital frailty risk score:*** We defined baseline frailty using the Hospital Frailty Risk Score defined by Gilbert et al.[14]. The Hospital Frailty Risk Score is a validated method of deriving a frailty score from routinely-collected inpatient data, using ICD-10 diagnosis codes with individual weightings. We grouped the frailty score into three levels (low: <5, medium: 5-15, and high: >15), where the score was calculated using diagnosis codes from all inpatient admissions in the APDC from the previous 24 months[15].

***Recent surgeries:*** We ascertained recent surgeries as per previous studies using the Medicines Intelligence Data Platform[16]. We defined recent surgeries using any inpatient hospital procedure with an ACHI surgical procedure code corresponding to ACHI code blocks identified in Gong et al.[17] in the 30 days prior to study opioid initiation. Defined as binary (1 = recent surgery, 0 = no recent surgery).

***History of falls:*** We ascertained previous falls in the 2 years to 61 days (inclusive) prior to study opioid initiation. We looked for evidence of falls resulting in ED presentation or hospitalisation (defined as in the outcome; Supplementary Table 4), defined as a binary indicator (1 = previous fall, 0 = no previous fall).

***Exposure to falls-risk medicines:*** We used an established list of Falls-Risk-Increasing Drugs (FRIDs) that has been used extensively in the literature and includes medicines known to increase the risk of falls and to cause orthostatism and hypotension[18, 19]. Defined as binary (1 = recent exposure to FRIDs, 0 = no recent exposure to FRIDs).

Covariates were ascertained as per the table below. Unless otherwise specified, diagnosis codes could occur in any position and any combination of codes present during the lookback window were sufficient to ascertain the condition or exposure of interest.

| Covariate | Data source | Codes |
| --- | --- | --- |
| Comorbidities | | |
| Cancer<br>(Dichotomous: 1 = yes, 0 = no) | NSW Cancer Registry | Between (initiation - 365) and (initiation - 1):<br>any record with diagnosis date within the ascertainment period |
| | ICD-10-AM code in EDDC or APDC | Between (initiation - 365) and (initiation - 1):<br>C00-C26, C30-C34, C37- C41, C43-C58, C60- C76, C81-C85, C88, C90-C97, Z51.1 |
| | ATC code in PBS | Between (initiation - 365) and (initiation - 1):<br>L01AA01-L01AX04, L01BA01, L01BA03- L01XX53, L02BG03, L02BG04, L02BG06, L02BB01-L02BB04, L02BX01-L02BX03, L04AX02, L04AX04, L04AX06, L02BA01, L02AE03, L02AE02 |
| History of substance use disorder<br>(Dichotomous: 1 = yes, 0 = no) | ICD-10-AM code in EDDC or APDC | Between (initiation - 365) and (initiation - 1):<br>F11, F12-F19, Z71.5, Z72.2 |
| | ATC code in PBS | Between (initiation - 365) and (initiation - 1):<br>N07B |
| History of overdose<br>(Dichotomous: 1 = yes, 0 = no) | ICD-10-AM code in EDDC or APDC | Between (initiation - 365) and (initiation - 1):<br>T40.0-T40.9, T42.3, T42.4, T42.6, T42.7, T43.6, T43.9, T51.0-T51.3, T51.8, T51.9, T52.0-T52.4, T52.8, T52.9, T53.0-T53.9 |
| Depression<br>(Dichotomous: 1 = yes, 0 = no) | ICD-10-AM code in EDDC or APDC | Between (initiation - 365) and (initiation - 1):<br>F20.4, F31.3-F31.5, F32, F33, F34.1, F41.2, F43.2, U79.3 |
| | ATC code in PBS | Between (initiation - 365) and (initiation - 1):<br>N06A |
| Anxiety<br>(Dichotomous: 1 = yes, 0 = no) | ICD-10-AM code in EDDC or APDC | Between (initiation - 365) and (initiation - 1):<br>F40-F41, F43.22, F43.23 |
| | ATC code in PBS | Between (initiation - 365) and (initiation - 1):<br>N05BA01–N05BA12, N05BE01 |

| Covariate | Data source | Codes |
| --- | --- | --- |
| Hypertension (Dichotomous: 1 = yes, 0 = no) | ICD-10-AM code in EDDC or APDC | Between (initiation - 365) and (initiation - 1): I10-I15, U82.3 |
| | ATC code in PBS | Between (initiation - 365) and (initiation - 1): C02AB01-C02AC05, C02DB02-C02KX01 |
| Atrial fibrillation / flutter (Dichotomous: 1 = yes, 0 = no) | ICD-10-AM code in EDDC or APDC | Between (initiation - 365) and (initiation - 1): I48 |
| Diabetes[20, 21] (Dichotomous: 1 = yes, 0 = no) | ICD-10-AM code in EDDC or APDC | Between (initiation - 365) and (initiation - 1): E10-E14 (excl E12) |
| | ATC code in PBS | Between (initiation - 365) and (initiation - 1): A10AA-A10AE, A10BA02, A10BD02, A10BD03, A10BD07, A10BD08, A10BD10, A10BD11, A10BD13, A10BB, A10BH, A10BD19, A10BD21, A10BD24, A10BG, A10BF |
| Congestive heart failure[21] (Dichotomous: 1 = yes, 0 = no) | ICD-10-AM code in EDDC or APDC | Between (initiation - 365) and (initiation - 1): I50 |
| | ATC code in PBS | Between (initiation - 365) and (initiation - 1): C03DA; C07AB02, C07AB07, C07AG02, C07AB12; (C03C) AND (C09A OR C09C) |
| Thyroid disease (Dichotomous: 1 = yes, 0 = no | ICD-10-AM code in EDDC or APDC | Between (initiation - 365) and (initiation - 1): E00-E07, E89.0 |
| | ATC code in PBS | Between (initiation - 365) and (initiation - 1): H03BA02 - H03BB01, H03AA01 - H03AA02 |
| Postural hypotension (Dichotomous: 1 = yes, 0 = no) | ICD-10-AM code in EDDC or APDC | Between (initiation - 365) and (initiation - 1): I95.1 |
| Osteoporosis[22] (Dichotomous: 1 = yes, 0 = no) | ICD-10-AM code in EDDC or APDC | Between (initiation - 365) and (initiation - 1): M80, M81, M82 |

| Covariate | Data source | Codes |
| --- | --- | --- |
| | ATC code in PBS | Between (initiation - 365) and (initiation - 1):<br>M05B |
| Stroke[23]<br>(Dichotomous: 1 = yes, 0 = no) | ICD-10-AM code in EDDC or APDC | Between (initiation - 365) and (initiation - 1):<br>I60, I61, I62, I64 |
| | ATC code in PBS | Between (initiation - 365) and (initiation - 1):<br>B01A |
| Renal impairment[24]<br>(Dichotomous: 1 = yes, 0 = no) | ICD-10-AM code in EDDC or APDC | Between (initiation - 365) and (initiation - 1):<br>I12.0, I13.1, N03.2 - N03.7, N05.2 - N05.7, N18, N19, N25.0, Z49.0 - Z49.2, Z94.0, Z99.2 |
| | ATC code in PBS | Between (initiation - 365) and (initiation - 1):<br>B03XA01-B03XA02, A11CC01-A11CC04, V03AE02 |
| Hepatic impairment[24]<br>(Dichotomous: 1 = yes, 0 = no) | ICD-10-AM code in EDDC or APDC | Between (initiation - 365) and (initiation - 1):<br>B18, K70.0 - K70.3, K70.9, K71.3 - K71.5, K71.7, K73, K74, K76.0, K76.2 - K76.4, K76.8, K76.9, Z94.4 |
| | ATC code in PBS | Between (initiation - 365) and (initiation - 1):<br>A06AD11 |
| Hospital Frailty Risk Score | | |
| Baseline frailty | ICD-10-AM code in APDC | Between (initiation - 730) and (initiation - 1):<br>A04, A09, A41, B95, B96, D64, E05, E16, E53, E55, E83, E86, E87, F00, F01, F03, F05, F10, F32, G20, G30, G31, G40, G45, G81, H54, H91, I63, I67, I69, I95, J18, J22, J69, J96, K26, K52, K59, K92, L03, L08, L89, L97, M15, M19, M25, M41, M48, M79, M80, M81, N17, N18, N19, N20, N28, N39, R00, R02, R11, R13, R26, R29, R31, R32, R33, R40, R41, R44, R45, R47, R50, R54, R55, R56, R63, R69, R79, R94, S00, S01, S06, S09, S22, S32, S42, S51, S72, S80, T83, U80, W01, W06, W10, W18, W19, X59, Y84, Y95, Z22, Z50, Z60, Z73, Z74, Z75, Z87, Z91, Z93, Z99 |
| Previous medicine use | | |

| Covariate | Data source | Codes |
|---|---|---|
| Recent opioid exposure<br>(Dichotomous: 1 = yes, 0 = no) | PBS | Between (initiation - 90) and (initiation - 1):<br>Record of dispensing with ATC N02AA, N02AA01, N02AA03, N02AA05 (except item codes for controlled-release oxycodone), N02AA59, N02AB02, N02AB03, N02AC, N02AC02, N02AC04, N02AE01, N02AJ06, N02AJ07, N02AX02 |
| Falls-risk medicines (FRIDs)[9]<br>(Dichotomous: 1 = yes, 0 = no) | PBS | Between (initiation - 30) and (initiation - 1):<br>Record of dispensing with ATC N05A (excluding N05AN), N05B, N05C, N06A, C01D, C02, C03, C07, C08, C09, G04CA, N04B |
| Gabapentinoids<br>(Dichotomous: 1 = yes, 0 = no) | PBS | Between (initiation - 30) and (initiation - 1):<br>Record of dispensing with ATC N03AX12, N03AX16, N02BG |
| Anticholinergic medicines<br>(Dichotomous: 1 = yes, 0 = no) | PBS | Between (initiation - 30) and (initiation - 1):<br>Record of dispensing with ATC N04A |
| Statins<br>(Dichotomous: 1 = yes, 0 = no) | PBS | Between (initiation - 30) and (initiation - 1):<br>Record of dispensing with ATC C10 |

APDC: Admitted Patient Data Collection; ATC: Anatomical Therapeutic Chemical classification; EDDC: Emergency Department Data Collection; ICD-10-AM: International Classification of Diseases, 10th Revision, Australian Modification; NSW: New South Wales; PBS: Pharmaceutical Benefits Scheme.

**Supplementary Table 6. Baseline characteristics of tapentadol (SR) and oxycodone (CR) initiators aged 65 years and older, 1 September 2014 - 3 December 2020, before and after propensity score matching**

| Characteristic | Unmatched | | | Matched | | |
|---|---|---|---|---|---|---|
| | Tapentadol SR N = 46,241 | Oxycodone CR N = 199,754 | SD | Tapentadol SR N = 41,546 | Oxycodone CR N = 41,546 | SD |
| Year of initiation* | | | | | | |
| 2014 | 1,284 (2.8%) | 12,795 (6.4%) | .17 | 1,212 (2.9%) | 1,212 (2.9%) | .00 |
| 2015 | 4,426 (9.6%) | 38,660 (19.4%) | .28 | 4,160 (10.0%) | 4,160 (10.0%) | .00 |
| 2016 | 5,163 (11.2%) | 39,484 (19.8%) | .24 | 4,894 (11.8%) | 4,894 (11.8%) | .00 |
| 2017 | 7,357 (15.9%) | 37,569 (18.8%) | .08 | 6,921 (16.7%) | 6,921 (16.7%) | .00 |
| 2018 | 8,622 (18.7%) | 30,539 (15.3%) | .09 | 7,999 (19.3%) | 7,999 (19.3%) | .00 |
| 2019 | 10,819 (23.4%) | 24,517 (12.3%) | .29 | 9,566 (23.0%) | 9,566 (23.0%) | .00 |
| 2020 | 8,570 (18.5%) | 16,190 (8.1%) | .31 | 6,794 (16.4%) | 6,794 (16.4%) | .00 |
| Sex | | | | | | |
| Female | 27,256 (58.9%) | 108,926 (54.5%) | .09 | 24,289 (58.5%) | 23,722 (57.1%) | .03 |
| Male | 18,985 (41.1%) | 90,828 (45.5%) | | 17,257 (41.5%) | 17,824 (42.9%) | |
| Age group (years) | | | | | | |
| 65-84 | 40,395 (87.4%) | 163,146 (81.7%) | .16 | 36,794 (88.6%) | 36,794 (88.6%) | .00 |
| 85 plus | 5,846 (12.6%) | 36,608 (18.3%) | .16 | 4,752 (11.4%) | 4,752 (11.4%) | .00 |
| Remoteness Area | | | | | | |
| Major cities | 29,699 (64.2%) | 126,846 (63.5%) | .02 | 26,620 (64.1%) | 26,626 (64.1%) | .00 |
| Inner regional | 13,084 (28.3%) | 54,875 (27.5%) | .02 | 11,796 (28.4%) | 11,609 (27.9%) | .01 |
| Outer regional | 3,072 (6.6%) | 15,963 (8.0%) | .05 | 2,786 (6.7%) | 2,924 (7.0%) | .01 |
| Remote | 160 (0.4%) | 996 (0.5%) | .02 | 139 (0.3%) | 165 (0.4%) | .01 |
| Very remote | 18 (0.0%) | 99 (0.1%) | .01 | 16 (0.0%) | 21 (0.1%) | .01 |
| Missing | 208 (0.5%) | 975 (0.5%) | .01 | 189 (0.5%) | 201 (0.5%) | .00 |
| Decile of relative disadvantage | | | | | | |
| 1 (most disadvantage) | 2,986 (6.5%) | 14,235 (7.1%) | .03 | 2,695 (6.5%) | 2,910 (7.0%) | .02 |
| 2 | 4,282 (9.3%) | 18,828 (9.4%) | .01 | 3,847 (9.3%) | 3,932 (9.5%) | .01 |
| 3 | 3,751 (8.1%) | 17,082 (8.6%) | .02 | 3,428 (8.3%) | 3,469 (8.4%) | .00 |

| Characteristic | Unmatched | | | Matched | | |
|---|---|---|---|---|---|---|
| | Tapentadol SR N = 46,241 | Oxycodone CR N = 199,754 | SD | Tapentadol SR N = 41,546 | Oxycodone CR N = 41,546 | SD |
| 4 | 5,568 (12.0%) | 25,638 (12.8%) | .02 | 5,070 (12.2%) | 5,074 (12.2%) | .00 |
| 5 | 6,117 (13.2%) | 23,761 (11.9%) | .04 | 5,286 (12.7%) | 5,323 (12.8%) | .00 |
| 6 | 5,061 (10.9%) | 21,853 (10.9%) | .00 | 4,585 (11.0%) | 4,594 (11.1%) | .00 |
| 7 | 4,371 (9.5%) | 18,879 (9.5%) | .00 | 3,945 (9.5%) | 4,051 (9.8%) | .01 |
| 8 | 4,034 (8.7%) | 17,967 (9.0%) | .01 | 3,635 (8.8%) | 3,661 (8.8%) | .00 |
| 9 | 5,467 (11.8%) | 21,974 (11.0%) | .03 | 4,911 (11.8%) | 4,616 (11.1%) | .02 |
| 10 (least disadvantage) | 4,386 (9.5%) | 18,533 (9.3%) | .01 | 3,946 (9.5%) | 3,703 (8.9%) | .02 |
| Missing | 218 (0.5%) | 1,004 (0.5%) | .00 | 198 (0.5%) | 213 (0.5%) | .01 |
| Exposure to non-study opioids (prior 90 days) | | | | | | |
| None | 29,940 (64.8%) | 117,517 (58.8%) | .12 | 27,056 (65.1%) | 27,056 (65.1%) | .00 |
| Yes | 16,301 (35.3%) | 82,237 (41.2%) | | 14,490 (34.9%) | 14,490 (34.9%) | |
| Comorbidities | | | | | | |
| Cancer | 5,537 (12.0%) | 39,155 (19.6%) | .21 | 4,596 (11.1%) | 4,864 (11.7%) | .02 |
| Substance use disorder | 1,184 (2.6%) | 5,977 (3.0%) | .03 | 1,032 (2.5%) | 1,214 (2.9%) | .03 |
| Prior overdose | 47 (0.1%) | 207 (0.1%) | .00 | 39 (0.1%) | 34 (0.1%) | .00 |
| Depression | 14,341 (31.0%) | 60,098 (30.1%) | .02 | 12,440 (29.9%) | 13,102 (31.5%) | .03 |
| Anxiety | 5,071 (11.0%) | 22,073 (11.1%) | .00 | 4,481 (10.8%) | 4,509 (10.9%) | .00 |
| Hypertension | 15,662 (33.9%) | 68,719 (34.4%) | .01 | 13,674 (32.9%) | 14,402 (34.7%) | .04 |
| Atrial fibrillation or flutter | 1,747 (3.8%) | 10,661 (5.3%) | .07 | 1,501 (3.6%) | 1,487 (3.6%) | .00 |
| Diabetes | 9,843 (21.3%) | 44,546 (22.3%) | .02 | 8,830 (21.3%) | 8,968 (21.6%) | .01 |
| Congestive heart failure | 10,930 (23.6%) | 52,451 (26.3%) | .06 | 9,635 (23.2%) | 9,741 (23.5%) | .01 |
| Thyroid disease | 5,539 (12.0%) | 22,403 (11.2%) | .02 | 4,881 (11.8%) | 4,792 (11.5%) | .01 |
| Postural hypotension | 414 (0.9%) | 2,352 (1.2%) | .03 | 344 (0.8%) | 388 (0.9%) | .01 |
| Osteoporosis | 10,284 (22.2%) | 51,043 (25.6%) | .08 | 8,917 (21.5%) | 8,859 (21.3%) | .00 |
| Stroke | 16,312 (35.3%) | 79,231 (39.7%) | .09 | 14,658 (35.3%) | 13,774 (33.2%) | .04 |
| Renal impairment | 1,269 (2.7%) | 8,614 (4.3%) | .09 | 1,071 (2.6%) | 1,099 (2.7%) | .00 |
| Hepatic impairment | 379 (0.8%) | 3,164 (1.6%) | .07 | 300 (0.7%) | 253 (0.6%) | .01 |

| Characteristic | Unmatched | | | Matched | | |
|---|---|---|---|---|---|---|
| | Tapentadol SR N = 46,241 | Oxycodone CR N = 199,754 | SD | Tapentadol SR N = 41,546 | Oxycodone CR N = 41,546 | SD |
| Surgery (prior 30 days) | 11,469 (24.8%) | 58,653 (29.4%) | .10 | 10,149 (24.4%) | 9,894 (23.8%) | .01 |
| Falls-risk medicines (FRIDs; prior 30 days) | | | | | | |
| Yes | 30,703 (66.4%) | 132,032 (66.1%) | .01 | 27,447 (66.1%) | 27,288 (65.7%) | .01 |
| Number of FRIDs: | | | | | | |
| 1 | 15,621 (50.9%) | 65,702 (49.8%) | .02 | 14,158 (51.6%) | 13,714 (50.3%) | .03 |
| 2 | 9,504 (31.0%) | 41,090 (31.1%) | .00 | 8,429 (30.7%) | 8,530 (31.3%) | .01 |
| 3 | 4,008 (13.1%) | 17,833 (13.5%) | .01 | 3,512 (12.8%) | 3,601 (13.2%) | .01 |
| 4 | 1,204 (3.9%) | 5,635 (4.3%) | .02 | 1,037 (3.8%) | 1,106 (4.1%) | .01 |
| 5+ | 366 (1.2%) | 1,772 (1.3%) | .01 | 311 (1.1%) | 337 (1.2%) | .01 |
| Type of FRID | | | | | | |
| *Medicines associated with high risk of falls* | | | | | | |
| Antipsychotics (excluding lithium) | 539 (1.8%) | 3,949 (3.0%) | .08 | 478 (1.7%) | 741 (2.7%) | .07 |
| Anxiolytics | 1,342 (4.4%) | 6,688 (5.1%) | .03 | 1,185 (4.3%) | 1,334 (4.9%) | .03 |
| Hypnotics and sedatives | 1,715 (5.6%) | 8,993 (6.8%) | .05 | 1,473 (5.4%) | 1,561 (5.7%) | .02 |
| Antidepressants | 8,395 (27.3%) | 36,337 (27.5%) | .00 | 7,248 (26.4%) | 7,949 (29.1%) | .06 |
| *Medicines that cause orthostatism/hypotension* | | | | | | |
| Vasodilators used in cardiac disease (e.g. nitrates) | 1,305 (4.3%) | 7,157 (5.4%) | .05 | 1,145 (4.2%) | 1,156 (4.2%) | .00 |
| Antihypertensives | 1,546 (5.0%) | 5,856 (4.4%) | .03 | 1,366 (5.0%) | 1,268 (4.7%) | .02 |
| Diuretics | 2,773 (9.0%) | 14,647 (11.1%) | .07 | 2,396 (8.7%) | 2,492 (9.1%) | .01 |
| Beta blockers | 5,909 (19.3%) | 26,537 (20.1%) | .02 | 5,269 (19.2%) | 5,177 (19.0%) | .01 |
| Calcium channel blockers | 6,616 (21.6%) | 28,879 (21.9%) | .01 | 5,895 (21.5%) | 5,869 (21.5%) | .00 |
| Renin-angiotensin acting agents | 21,205 (69.1%) | 85,846 (65.0%) | .09 | 19,062 (69.5%) | 18,473 (67.7%) | .04 |
| Alpha adrenoreceptor blockers | 1,058 (3.5%) | 4,409 (3.3%) | .01 | 943 (3.4%) | 924 (3.4%) | .00 |
| Dopaminergic agents | 969 (3.2%) | 3,845 (2.9%) | .01 | 854 (3.1%) | 799 (2.9%) | .01 |
| Other medicines associated with falls (prior 30 days) | | | | | | |
| Gabapentinoids | 5,923 (12.8%) | 19,045 (9.5%) | .10 | 4,406 (10.6%) | 4,203 (10.1%) | .02 |
| Anticholinergic medicines | 14 (0.0%) | 75 (0.0%) | .00 | 12 (0.0%) | 10 (0.0%) | .00 |

| Characteristic | Unmatched | | | Matched | | |
|---|---|---|---|---|---|---|
| | Tapentadol SR N = 46,241 | Oxycodone CR N = 199,754 | SD | Tapentadol SR N = 41,546 | Oxycodone CR N = 41,546 | SD |
| Statins | 19,430 (42.0%) | 78,596 (39.4%) | .05 | 17,393 (41.9%) | 17,064 (41.1%) | .02 |
| Hospital frailty risk score | | | | | | |
| Low: <5 | 38,109 (82.4%) | 150,135 (75.2%) | .18 | 34,536 (83.1%) | 34,251 (82.4%) | .02 |
| Medium: 5-15 | 5,792 (12.5%) | 33,224 (16.6%) | .12 | 5,101 (12.3%) | 5,140 (12.4%) | .00 |
| High: >15 | 2,340 (5.1%) | 16,395 (8.2%) | .13 | 1,909 (4.6%) | 2,155 (5.2%) | .03 |
| History of falls | | | | | | |
| Falls in the past 2 years to 60 days | 2,792 (6.0%) | 16,826 (8.4%) | .09 | 2,397 (5.8%) | 2,657 (6.4%) | .03 |

Note: cohort matched on propensity score, year and month of initiation, age, and exposure to non-study opioids in the prior 90 days

* study period: 01 September 2014 – 03 December 2020

SR = sustained release; CR = controlled release; SD = absolute standardised difference

**Supplementary Table 7. Baseline characteristics of tapentadol (SR) and oxycodone (CR) initiators aged 80 years and older, 1 September 2014 - 3 December 2020, before and after propensity score matching**

| Characteristic | Unmatched | | | Matched | | |
|---|---|---|---|---|---|---|
| | Tapentadol SR N = 12,563 | Oxycodone CR N = 67,439 | SD | Tapentadol SR N = 10,783 | Oxycodone CR N = 10,783 | SD |
| Year of initiation* | | | | | | |
| 2014 | 393 (3.1%) | 4,225 (6.3%) | .15 | 360 (3.3%) | 360 (3.3%) | .00 |
| 2015 | 1,362 (10.8%) | 12,959 (19.2%) | .24 | 1,247 (11.6%) | 1,247 (11.6%) | .00 |
| 2016 | 1,518 (12.1%) | 13,106 (19.4%) | .20 | 1,390 (12.9%) | 1,390 (12.9%) | .00 |
| 2017 | 2,040 (16.2%) | 12,383 (18.4%) | .06 | 1,842 (17.1%) | 1,842 (17.1%) | .00 |
| 2018 | 2,260 (18.0%) | 10,316 (15.3%) | .07 | 2,000 (18.6%) | 2,000 (18.6%) | .00 |
| 2019 | 2,663 (21.2%) | 8,471 (12.6%) | .23 | 2,224 (20.6%) | 2,224 (20.6%) | .00 |
| 2020 | 2,327 (18.5%) | 5,979 (8.9%) | .28 | 1,720 (16.0%) | 1,720 (16.0%) | .00 |
| Sex | | | | | | |
| Female | 7,905 (62.92%) | 40,828 (60.5%) | .05 | 6,759 (62.7%) | 6,786 (62.9%) | .01 |
| Male | 4,658 (37.08%) | 26,611 (39.5%) | | 4,024 (37.3%) | 3,997 (37.1%) | |
| Age group (years) | | | | | | |
| 65-84 | 6,717 (53.5%) | 30,831 (45.7%) | .16 | 6,031 (55.9%) | 6,031 (55.9%) | .00 |
| 85 plus | 5,846 (46.5%) | 36,608 (54.3%) | .16 | 4,752 (44.1%) | 4,752 (44.1%) | |
| Remoteness Area | | | | | | |
| Major cities | 8,396 (66.8%) | 43,736 (64.9%) | .04 | 7,170 (66.5%) | 7,064 (65.5%) | .02 |
| Inner regional | 3,354 (26.7%) | 18,049 (26.8%) | .00 | 2,906 (27.0%) | 2,923 (27.1%) | .00 |
| Outer regional | 741 (5.9%) | 5,032 (7.5%) | .06 | 645 (6.0%) | 713 (6.6%) | .03 |
| Remote / Very Remote† | 40 (0.3%) | 328 (0.5%) | .03 | 34 (0.3%) | 42 (0.4%) | .01 |
| Missing | 32 (0.3%) | 294 (0.4%) | .03 | 28 (0.3%) | 41 (0.4%) | .02 |
| Decile of relative disadvantage | | | | | | |
| 1 (most disadvantage) | 783 (6.2%) | 4,524 (6.7%) | .02 | 676 (6.3%) | 742 (6.9%) | .02 |
| 2 | 1,168 (9.3%) | 6,282 (9.3%) | .00 | 1,000 (9.3%) | 1,039 (9.6%) | .01 |
| 3 | 922 (7.3%) | 5,458 (8.1%) | .03 | 817 (7.6%) | 845 (7.8%) | .01 |
| 4 | 1,381 (11.0%) | 8,250 (12.2%) | .04 | 1,199 (11.1%) | 1,257 (11.7%) | .02 |

| Characteristic | Unmatched | | | Matched | | |
|---|---|---|---|---|---|---|
| | Tapentadol SR<br>N = 12,563 | Oxycodone CR<br>N = 67,439 | SD | Tapentadol SR<br>N = 10,783 | Oxycodone CR<br>N = 10,783 | SD |
| 5 | 1,761 (14.0%) | 8,533 (12.7%) | .04 | 1,471 (13.6%) | 1,415 (13.1%) | .02 |
| 6 | 1,376 (11.0%) | 7,355 (10.9%) | .00 | 1,203 (11.2%) | 1,152 (10.7%) | .02 |
| 7 | 1,158 (9.2%) | 6,158 (9.1%) | .00 | 998 (9.3%) | 1,013 (9.4%) | .00 |
| 8 | 1,143 (9.1%) | 6,237 (9.3%) | .01 | 987 (9.2%) | 1,003 (9.3%) | .01 |
| 9 | 1,512 (12.0%) | 7,660 (11.4%) | .02 | 1,285 (11.9%) | 1,266 (11.7%) | .01 |
| 10 (least disadvantage) | 1,324 (10.5%) | 6,680 (9.9%) | .02 | 1,116 (10.4%) | 1,008 (9.4%) | .03 |
| Missing | 35 (0.3%) | 302 (0.5%) | .03 | 31 (0.3%) | 43 (0.4%) | .02 |
| Exposure to non-study opioids (prior 90 days) | | | | | | |
| None | 7,802 (62.1%) | 38,428 (57.0%) | .10 | 6,812 (63.2%) | 6,812 (63.2%) | .00 |
| Yes | 4,761 (37.9%) | 29,011 (43.0%) | | 3,971 (36.8%) | 3,971 (36.8%) | |
| Comorbidities | | | | | | |
| Cancer | 1,621 (12.9%) | 12,707 (18.8%) | .16 | 1,233 (11.4%) | 1,256 (11.7%) | .01 |
| Substance use disorder | 144 (1.2%) | 1,020 (1.5%) | .03 | 116 (1.1%) | 167 (1.6%) | .04 |
| Prior overdose | 19 (0.2%) | 76 (0.1%) | .01 | 16 (0.2%) | 15 (0.1%) | .00 |
| Depression | 3,928 (31.3%) | 21,341 (31.6%) | .01 | 3,274 (30.4%) | 3,572 (33.1%) | .06 |
| Anxiety | 1,334 (10.6%) | 7,295 (10.8%) | .01 | 1,138 (10.6%) | 1,150 (10.7%) | .00 |
| Hypertension | 4,763 (37.9%) | 25,732 (38.2%) | .01 | 3,981 (36.9%) | 4,194 (38.9%) | .04 |
| Atrial fibrillation or flutter | 746 (5.9%) | 5,182 (7.7%) | .07 | 610 (5.7%) | 597 (5.5%) | .01 |
| Diabetes | 2,470 (19.7%) | 13,769 (20.4%) | .02 | 2,137 (19.8%) | 2,209 (20.5%) | .02 |
| Congestive heart failure | 4,286 (34.1%) | 24,693 (36.6%) | .05 | 3,616 (33.5%) | 3,674 (34.1%) | .01 |
| Thyroid disease | 1,715 (13.7%) | 8,748 (13.0%) | .02 | 1,461 (13.6%) | 1,474 (13.7%) | .00 |
| Postural hypotension | 218 (1.7%) | 1,334 (2.0%) | .02 | 180 (1.7%) | 177 (1.6%) | .00 |
| Osteoporosis | 3,999 (31.8%) | 22,230 (33.0%) | .02 | 3,338 (31.0%) | 3,305 (30.7%) | .01 |
| Stroke | 5,817 (46.3%) | 32,893 (48.8%) | .05 | 4,991 (46.3%) | 4,699 (43.6%) | .05 |
| Renal impairment | 593 (4.7%) | 4,620 (6.9%) | .09 | 475 (4.4%) | 491 (4.6%) | .01 |
| Hepatic impairment | 112 (0.9%) | 1,114 (1.7%) | .07 | 90 (0.8%) | 69 (0.6%) | .02 |
| Surgery (prior 30 days) | 2,694 (21.4%) | 16,790 (24.9%) | .08 | 2,249 (20.9%) | 2,171 (20.1%) | .02 |

| Characteristic | Unmatched | | | Matched | | |
|---|---|---|---|---|---|---|
| | Tapentadol SR N = 12,563 | Oxycodone CR N = 67,439 | SD | Tapentadol SR N = 10,783 | Oxycodone CR N = 10,783 | SD |
| Falls-risk medicines (FRIDs; prior 30 days) | | | | | | |
| Yes | 9,216 (73.4%) | 48,761 (72.3%) | .02 | 7,889 (73.2%) | 7,863 (72.9%) | .01 |
| Number of FRIDs: | | | | | | |
| 1 | 4,228 (45.9%) | 21,897 (44.9%) | .02 | 3,681 (46.7%) | 3,406 (43.3%) | .07 |
| 2 | 2,978 (32.3%) | 15,778 (32.4%) | .00 | 2,508 (31.8%) | 2,668 (33.9%) | .05 |
| 3 | 1,426 (15.5%) | 7,584 (15.6%) | .00 | 1,207 (15.3%) | 1,240 (15.8%) | .01 |
| 4 | 455 (4.9%) | 2,645 (5.4%) | .02 | 383 (4.9%) | 412 (5.2%) | .02 |
| 5+ | 129 (1.4%) | 857 (1.8%) | .03 | 110 (1.4%) | 137 (1.7%) | .03 |
| Type of FRID | | | | | | |
| *Medicines associated with high risk of falls* | | | | | | |
| Antipsychotics (excluding lithium) | 180 (2.0%) | 1,910 (3.9%) | .12 | 154 (2.0%) | 266 (3.4%) | .09 |
| Anxiolytics | 382 (4.1%) | 2,293 (4.7%) | .03 | 325 (4.1%) | 342 (4.4%) | .01 |
| Hypnotics and sedatives | 659 (7.2%) | 4,328 (8.9%) | .06 | 542 (6.9%) | 614 (7.8%) | .04 |
| Antidepressants | 2,230 (24.2%) | 12,963 (26.6%) | .05 | 1,848 (23.4%) | 2,156 (27.4%) | .09 |
| *Medicines that cause orthostatism/hypotension* | | | | | | |
| Vasodilators used in cardiac disease (e.g. nitrates) | 659 (7.2%) | 4,087 (8.4%) | .05 | 553 (7.0%) | 556 (7.1%) | .00 |
| Antihypertensives | 424 (4.6%) | 1,990 (4.1%) | .03 | 358 (4.5%) | 323 (4.1%) | .02 |
| Diuretics | 1,232 (13.4%) | 7,852 (16.1%) | .08 | 1,016 (12.9%) | 1,092 (13.9%) | .03 |
| Beta blockers | 2,110 (22.9%) | 11,210 (23.0%) | .00 | 1,804 (22.9%) | 1,805 (23.0%) | .00 |
| Calcium channel blockers | 2,329 (25.3%) | 11,938 (24.5%) | .02 | 1,995 (25.3%) | 2,041 (26.0%) | .02 |
| Renin-angiotensin acting agents | 6,010 (65.2%) | 29,035 (59.6%) | .12 | 5,200 (65.9%) | 4,998 (63.6%) | .05 |
| Alpha adrenoreceptor blockers | 405 (4.4%) | 1,990 (4.1%) | .02 | 341 (4.3%) | 352 (4.5%) | .01 |
| Dopaminergic agents | 332 (3.6%) | 1,660 (3.4%) | .01 | 281 (3.6%) | 274 (3.5%) | .00 |
| Other medicines associated with falls (prior 30 days) | | | | | | |
| Gabapentinoids | 1,625 (12.9%) | 6,358 (9.4%) | .11 | 1,119 (10.4%) | 1,010 (9.4%) | .03 |
| Anticholinergic medicines | 6 (0.1%) | 13 (0.0%) | .02 | <6 | <6 | ** |
| Statins | 5,526 (44.0%) | 26,226 (38.9%) | .10 | 4,763 (44.2%) | 4,559 (42.3%) | .04 |
| Hospital frailty risk score | | | | | | |

| Characteristic | Unmatched | | | Matched | | |
|---|---|---|---|---|---|---|
| | **Tapentadol SR** | **Oxycodone CR** | **SD** | **Tapentadol SR** | **Oxycodone CR** | **SD** |
| | **N = 12,563** | **N = 67,439** | | **N = 10,783** | **N = 10,783** | |
| Low: <5 | 8,989 (71.6%) | 42,406 (62.9%) | .19 | 7,834 (72.7%) | 7,689 (71.3%) | .03 |
| Medium: 5-15 | 2,296 (18.3%) | 14,786 (21.9%) | .09 | 1,958 (18.2%) | 1,899 (17.6%) | .01 |
| High: >15 | 1,278 (10.2%) | 10,247 (15.2%) | .15 | 991 (9.2%) | 1,195 (11.1%) | .06 |
| History of falls | | | | | | |
| Falls in the past 2 years to 60 days | 1,468 (11.7%) | 10,520 (15.6%) | .11 | 1,206 (11.2%) | 1,389 (12.9%) | .05 |

Note: cohort matched on propensity score, year and month of initiation, age, and exposure to non-study opioids in the prior 90 days

SR = sustained release; CR = controlled release; SD = absolute standardised difference

* study period: 01 September 2014 – 03 December 2020

** suppressed to avoid residual disclosure of small cells

† categories combined to avoid residual disclosure of small cells

**Supplementary Table 8. Unadjusted relative risks of falls among people initiating tapentadol (SR) compared to oxycodone (CR), overall and by recent exposure to non-study opioids, for the total study population, people aged 65+, and people aged 80+**

| Outcome | Exposure | Overall | | | No recent exposure | | | Exposure in prior 90 days | | |
|---|---|---|---|---|---|---|---|---|---|---|
| | | No. exposed | No. events | RR (95% CI) | No. exposed | No. events | RR (95% CI) | No. exposed | No. events | RR (95% CI) |
| Overall population | | | | | | | | | | |
| 7 days post-initiation | Oxycodone CR | 419,732 | 1,396 | 0.55 (0.47-0.64) | 250,030 | 748 | 0.61 (0.50-0.73) | 169,702 | 648 | 0.49 (0.38-0.62) |
| | Tapentadol SR | 103,924 | 190 | | 65,715 | 119 | | 38,209 | 71 | |
| 14 days post-initiation | Oxycodone CR | 419,732 | 2,108 | 0.58 (0.52-0.66) | 250,030 | 1,147 | 0.60 (0.52-0.71) | 169,702 | 961 | 0.56 (0.47-0.68) |
| | Tapentadol SR | 103,924 | 304 | | 65,715 | 182 | | 38,209 | 122 | |
| 28 days post-initiation | Oxycodone CR | 419,732 | 3,007 | 0.62 (0.56-0.68) | 250,030 | 1,618 | 0.63 (0.56-0.72) | 169,702 | 1,389 | 0.61 (0.52-0.71) |
| | Tapentadol SR | 103,924 | 460 | | 65,715 | 270 | | 38,209 | 190 | |
| People aged 65 years and older | | | | | | | | | | |
| 7 days post-initiation | Oxycodone CR | 199,754 | 1,128 | 0.58 (0.49-0.69) | 117,517 | 609 | 0.61 (0.49-0.76) | 82,237 | 519 | 0.55 (0.42-0.73) |
| | Tapentadol SR | 46,241 | 152 | | 29,940 | 95 | | 16,301 | 57 | |
| 14 days post-initiation | Oxycodone CR | 199,754 | 1,723 | 0.60 (0.53-0.69) | 117,517 | 942 | 0.60 (0.51-0.72) | 82,237 | 781 | 0.61 (0.50-0.76) |
| | Tapentadol SR | 46,241 | 240 | | 29,940 | 145 | | 16,301 | 95 | |
| 28 days post-initiation | Oxycodone CR | 199,754 | 2,480 | 0.63 (0.57-0.71) | 117,517 | 1,354 | 0.63 (0.55-0.73) | 82,237 | 1,126 | 0.66 (0.56-0.78) |
| | Tapentadol SR | 46,241 | 364 | | 29,940 | 217 | | 16,301 | 147 | |
| People aged 80 years and older | | | | | | | | | | |
| 7 days post-initiation | Oxycodone CR | 67,439 | 701 | 0.71 (0.57-0.88) | 38,428 | 403 | 0.71 (0.54-0.93) | 29,011 | 298 | 0.72 (0.50-1.01) |
| | Tapentadol SR | 12,563 | 93 | | 7,802 | 58 | | 4,761 | 35 | |
| 14 days post-initiation | Oxycodone CR | 67,439 | 1,091 | 0.73 (0.62-0.87) | 38,428 | 631 | 0.73 (0.58-0.90) | 29,011 | 460 | 0.74 (0.56-0.98) |
| | Tapentadol SR | 12,563 | 149 | | 7,802 | 93 | | 4,761 | 56 | |
| 28 days post-initiation | Oxycodone CR | 67,439 | 1,606 | 0.75 (0.65-0.86) | 38,428 | 922 | 0.74 (0.62-0.89) | 29,011 | 684 | 0.76 (0.61-0.95) |
| | Tapentadol SR | 12,563 | 224 | | 7,802 | 139 | | 4,761 | 85 | |

SR = sustained release; CR = controlled release; RR = relative risk; CI = confidence interval

**Supplementary Table 9. Baseline characteristics of tapentadol (SR) and oxycodone (CR) initiators with only study opioids dispensed on initiation date, 1 September 2014 - 3 December 2020, before and after propensity score matching**

| Characteristic | Unmatched | | | Matched | | |
|---|---|---|---|---|---|---|
| | Tapentadol SR N = 87,542 | Oxycodone CR N = 281,697 | SD | Tapentadol SR N = 86,804 | Oxycodone CR N = 86,804 | SD |
| Year of initiation* | | | | | | |
| 2014 | 2,556 (2.9%) | 17,305 (6.1%) | .16 | 2,554 (2.9%) | 2,554 (2.9%) | .00 |
| 2015 | 8,602 (9.8%) | 53,326 (18.9%) | .26 | 8,589 (9.9%) | 8,589 (9.9%) | .00 |
| 2016 | 9,826 (11.2%) | 55,837 (19.8%) | .24 | 9,813 (11.3%) | 9,813 (11.3%) | .00 |
| 2017 | 14,470 (16.5%) | 54,180 (19.2%) | .07 | 14,454 (16.7%) | 14,454 (16.7%) | .00 |
| 2018 | 16,177 (18.5%) | 44,268 (15.7%) | .07 | 16,138 (18.6%) | 16,138 (18.6%) | .00 |
| 2019 | 20,057 (22.9%) | 34,603 (12.3%) | .28 | 19,932 (23.0%) | 19,932 (23.0%) | .00 |
| 2020 | 15,854 (18.1%) | 22,178 (7.9%) | .31 | 15,324 (17.7%) | 15,324 (17.7%) | .00 |
| Sex | | | | | | |
| Female | 51,214 (58.5%) | 155,652 (55.3%) | .07 | 50,702 (58.4%) | 50,770 (58.5%) | .00 |
| Male | 36,328 (41.5%) | 126,045 (44.7%) | | 36,102 (41.6%) | 36,034 (41.5%) | |
| Age group (years) | | | | | | |
| 18-44 | 17,022 (19.4%) | 53,811 (19.1%) | .01 | 16,954 (19.5%) | 17,234 (19.9%) | .01 |
| 45-64 | 30,435 (34.8%) | 85,885 (30.5%) | .09 | 30,092 (34.7%) | 29,937 (34.5%) | .00 |
| 65-84 | 34,711 (39.7%) | 111,868 (39.7%) | .00 | 34,414 (39.7%) | 34,123 (39.3%) | .01 |
| 85 plus | 5,374 (6.1%) | 30,133 (10.7%) | .16 | 5,344 (6.2%) | 5,510 (6.4%) | .01 |
| Remoteness Area | | | | | | |
| Major cities | 57,890 (66.1%) | 184,394 (65.5%) | .01 | 57,422 (66.2%) | 57,580 (66.3%) | .00 |
| Inner regional | 23,192 (26.5%) | 72,110 (25.6%) | .02 | 22,950 (26.4%) | 22,819 (26.3%) | .00 |
| Outer regional | 5,690 (6.5%) | 21,958 (7.8%) | .05 | 5,665 (6.5%) | 5,623 (6.5%) | .00 |
| Remote | 354 (0.4%) | 1,752 (0.6%) | .03 | 353 (0.4%) | 373 (0.4%) | .00 |
| Very remote | 50 (0.1%) | 195 (0.1%) | .00 | 49 (0.1%) | 54 (0.1%) | .00 |
| Missing | 366 (0.4%) | 1,288 (0.5%) | .01 | 365 (0.4%) | 355 (0.4%) | .00 |
| Decile of relative disadvantage | | | | | | |
| 1 (most disadvantage) | 6,527 (7.5%) | 22,747 (8.1%) | .02 | 6,495 (7.5%) | 6,255 (7.2%) | .01 |

| Characteristic | Unmatched | | | Matched | | |
|---|---|---|---|---|---|---|
| | Tapentadol SR N = 87,542 | Oxycodone CR N = 281,697 | SD | Tapentadol SR N = 86,804 | Oxycodone CR N = 86,804 | SD |
| 2 | 8,073 (9.2%) | 26,290 (9.3%) | .00 | 8,006 (9.2%) | 8,067 (9.3%) | .00 |
| 3 | 7,101 (8.1%) | 23,665 (8.4%) | .01 | 7,059 (8.1%) | 6,917 (8.0%) | .01 |
| 4 | 10,366 (11.8%) | 35,127 (12.5%) | .02 | 10,294 (11.9%) | 10,373 (12.0%) | .00 |
| 5 | 11,503 (13.1%) | 31,621 (11.2%) | .06 | 11,312 (13.0%) | 11,268 (13.0%) | .00 |
| 6 | 9,929 (11.3%) | 31,952 (11.3%) | .00 | 9,850 (11.4%) | 9,828 (11.3%) | .00 |
| 7 | 8,833 (10.1%) | 28,277 (10.0%) | .00 | 8,752 (10.1%) | 8,907 (10.3%) | .01 |
| 8 | 7,639 (8.7%) | 25,039 (8.9%) | .01 | 7,593 (8.8%) | 7,698 (8.9%) | .00 |
| 9 | 9,651 (11.0%) | 30,703 (10.9%) | .00 | 9,580 (11.0%) | 9,623 (11.1%) | .00 |
| 10 (least disadvantage) | 7,533 (8.6%) | 24,944 (8.9%) | .01 | 7,478 (8.6%) | 7,500 (8.6%) | .00 |
| Missing | 387 (0.4%) | 1,332 (0.5%) | .00 | 385 (0.4%) | 368 (0.4%) | .00 |
| Exposure to non-study opioids (prior 90 days) | | | | | | |
| None | 56,947 (65.1%) | 171,359 (60.8%) | .09 | 56,293 (64.9%) | 56,293 (64.9%) | .00 |
| Yes | 30,595 (35.0%) | 110,338 (39.2%) | | 30,511 (35.2%) | 30,511 (35.2%) | |
| Comorbidities | | | | | | |
| Cancer | 7,541 (8.6%) | 39,665 (14.1%) | .17 | 7,506 (8.7%) | 7,554 (8.7%) | .00 |
| Substance use disorder | 3,868 (4.4%) | 12,910 (4.6%) | .01 | 3,845 (4.4%) | 3,860 (4.5%) | .00 |
| Prior overdose | 209 (0.2%) | 671 (0.2%) | .00 | 206 (0.2%) | 202 (0.2%) | .00 |
| Depression | 30,473 (34.8%) | 88,373 (31.4%) | .07 | 30,118 (34.7%) | 30,028 (34.6%) | .00 |
| Anxiety | 11,731 (13.4%) | 36,247 (12.9%) | .02 | 11,623 (13.4%) | 11,367 (13.1%) | .01 |
| Hypertension | 18,322 (20.9%) | 61,777 (21.9%) | .02 | 18,073 (20.8%) | 19,193 (22.1%) | .03 |
| Atrial fibrillation or flutter | 1,843 (2.1%) | 9,039 (3.2%) | .07 | 1,831 (2.1%) | 1,847 (2.1%) | .00 |
| Diabetes | 13,991 (16.0%) | 48,474 (17.2%) | .03 | 13,879 (16.0%) | 14,068 (16.2%) | .01 |
| Congestive heart failure | 12,419 (14.2%) | 47,942 (17.0%) | .08 | 12,323 (14.2%) | 12,421 (14.3%) | .00 |
| Thyroid disease | 7,783 (8.9%) | 24,376 (8.7%) | .01 | 7,695 (8.9%) | 7,648 (8.8%) | .00 |
| Postural hypotension | 458 (0.5%) | 2,011 (0.7%) | .02 | 452 (0.5%) | 478 (0.6%) | .00 |
| Osteoporosis | 11,189 (12.8%) | 46,096 (16.4%) | .10 | 11,109 (12.8%) | 11,211 (12.9%) | .00 |
| Stroke | 18,511 (21.2%) | 69,778 (24.8%) | .09 | 18,382 (21.2%) | 17,650 (20.3%) | .02 |
| Renal impairment | 1,444 (1.7%) | 7,714 (2.7%) | .07 | 1,427 (1.6%) | 1,359 (1.6%) | .01 |

| Characteristic | Unmatched | | | Matched | | |
|---|---|---|---|---|---|---|
| | Tapentadol SR<br>N = 87,542 | Oxycodone CR<br>N = 281,697 | SD | Tapentadol SR<br>N = 86,804 | Oxycodone CR<br>N = 86,804 | SD |
| Hepatic impairment | 816 (0.9%) | 4,767 (1.7%) | .07 | 811 (0.9%) | 648 (0.8%) | .02 |
| Surgery (prior 30 days) | 16,511 (18.9%) | 62,729 (22.3%) | .08 | 16,354 (18.8%) | 16,494 (19.0%) | .00 |
| Falls-risk medicines (FRIDs; prior 30 days) | | | | | | |
| Yes | 45,121 (51.5%) | 145,547 (51.7%) | .00 | 44,681 (51.5%) | 44,445 (51.2%) | .01 |
| Number of FRIDs: | | | | | | |
| 1 | 25,863 (57.3%) | 79,853 (54.9%) | .05 | 25,616 (57.3%) | 24,746 (55.7%) | .03 |
| 2 | 12,712 (28.2%) | 42,056 (28.9%) | .02 | 12,584 (28.2%) | 12,816 (28.8%) | .01 |
| 3 | 4,817 (10.7%) | 16,830 (11.6%) | .03 | 4,762 (10.7%) | 4,913 (11.1%) | .01 |
| 4 | 1,329 (3.0%) | 5,221 (3.6%) | .04 | 1,321 (3.0%) | 1,519 (3.4%) | .03 |
| 5+ | 400 (0.9%) | 1,587 (1.1%) | .02 | 398 (0.9%) | 451 (1.0%) | .01 |
| Type of FRID | | | | | | |
| *Medicines associated with high risk of falls* | | | | | | |
| Antipsychotics (excluding lithium) | 1,649 (3.7%) | 6,561 (4.5%) | .04 | 1,635 (3.7%) | 1,937 (4.4%) | .04 |
| Anxiolytics | 3,521 (7.8%) | 11,917 (8.2%) | .01 | 3,496 (7.8%) | 3,703 (8.3%) | .02 |
| Hypnotics and sedatives | 2,377 (5.3%) | 9,370 (6.4%) | .05 | 2,362 (5.3%) | 2,401 (5.4%) | .01 |
| Antidepressants | 17,230 (38.2%) | 51,403 (35.3%) | .06 | 17,021 (38.1%) | 17,752 (39.9%) | .04 |
| *Medicines that cause orthostatism/hypotension* | | | | | | |
| Vasodilators used in cardiac disease (e.g. nitrates) | 1,357 (3.0%) | 6,241 (4.3%) | .07 | 1,350 (3.0%) | 1,460 (3.3%) | .02 |
| Antihypertensives | 1,835 (4.1%) | 5,516 (3.8%) | .01 | 1,810 (4.1%) | 1,740 (3.9%) | .01 |
| Diuretics | 3,007 (6.7%) | 13,072 (9.0%) | .09 | 2,990 (6.7%) | 3,262 (7.3%) | .03 |
| Beta blockers | 6,873 (15.2%) | 24,736 (17.0%) | .05 | 6,829 (15.3%) | 6,896 (15.5%) | .01 |
| Calcium channel blockers | 7,148 (15.8%) | 25,234 (17.3%) | .04 | 7,073 (15.8%) | 7,169 (16.1%) | .01 |
| Renin-angiotensin acting agents | 25,984 (57.6%) | 82,637 (56.8%) | .02 | 25,721 (57.6%) | 25,226 (56.8%) | .02 |
| Alpha adrenoreceptor blockers | 1,033 (2.3%) | 3,424 (2.4%) | .00 | 1,025 (2.3%) | 935 (2.1%) | .01 |
| Dopaminergic agents | 1,121 (2.5%) | 3,491 (2.4%) | .01 | 1,113 (2.5%) | 1,065 (2.4%) | .01 |
| Other medicines associated with falls (prior 30 days) | | | | | | |
| Gabapentinoids | 10,344 (11.8%) | 23,314 (8.3%) | .12 | 10,060 (11.6%) | 9,923 (11.4%) | .00 |

| Characteristic | Unmatched | | | Matched | | |
|---|---|---|---|---|---|---|
| | **Tapentadol SR** | **Oxycodone CR** | **SD** | **Tapentadol SR** | **Oxycodone CR** | **SD** |
| | **N = 87,542** | **N = 281,697** | | **N = 86,804** | **N = 86,804** | |
| Anticholinergic medicines | 26 (0.0%) | 133 (0.1%) | .01 | 26 (0.0%) | 17 (0.0%) | .01 |
| Statins | 22,948 (26.2%) | 73,153 (26.0%) | .01 | 22,689 (26.1%) | 22,701 (26.2%) | .00 |
| Hospital frailty risk score | | | | | | |
| Low: <5 | 77,397 (88.4%) | 233,949 (83.1%) | .15 | 76,715 (88.4%) | 76,708 (88.4%) | .00 |
| Medium: 5-15 | 7,584 (8.7%) | 33,076 (11.7%) | .10 | 7,553 (8.7%) | 7,486 (8.6%) | .00 |
| High: >15 | 2,561 (2.9%) | 14,672 (5.2%) | .12 | 2,536 (2.9%) | 2,610 (3.0%) | .01 |
| History of falls | | | | | | |
| Falls in the past 2 years to 60 days | 3,374 (3.9%) | 15,838 (5.6%) | .08 | 3,347 (3.9%) | 3,475 (4.0%) | .01 |

Note: cohort matched on propensity score, year and month of initiation, and recent exposure to opioids

* study period: 01 September 2014 – 03 December 2020

SR = sustained release; CR = controlled release; SD = absolute standardised difference

**Supplementary Table 10. Relative risks of falls and cataract surgeries among tapentadol (SR) and oxycodone (CR) initiators with only study opioids dispensed on initiation date**

| Outcome | Exposure | Unmatched | | | Matched | | |
|---|---|---|---|---|---|---|---|
| | | No. exposed | No. events | RR (95% CI) | No. exposed | No. events | Matched RR[1] (95% CI) |
| Falls resulting in ED presentation, hospitalisation, or death | | | | | | | |
| 7 days post-initiation | Oxycodone CR | 281,697 | 957 | 0.56 (0.47-0.66) | 86,804 | 263 | 0.62 (0.51-0.75) |
| | Tapentadol SR | 87,542 | 166 | | 86,804 | 163 | |
| 14 days post-initiation | Oxycodone CR | 281,697 | 1,485 | 0.56 (0.49-0.64) | 86,804 | 414 | 0.62 (0.53-0.72) |
| | Tapentadol SR | 87,542 | 260 | | 86,804 | 257 | |
| 28 days post-initiation | Oxycodone CR | 281,697 | 2,162 | 0.59 (0.53-0.65) | 86,804 | 585 | 0.67 (0.59-0.76) |
| | Tapentadol SR | 87,542 | 395 | | 86,804 | 390 | |
| Cataract surgeries (negative control outcome) | | | | | | | |
| 7 days post-initiation | Oxycodone CR | 281,697 | 96 | 0.97 (0.64-1.47) | 86,804 | 22 | 1.32 (0.76-2.29) |
| | Tapentadol SR | 87,542 | 29 | | 86,804 | 29 | |
| 14 days post-initiation | Oxycodone CR | 281,697 | 201 | 1.12 (0.85-1.47) | 86,804 | 63 | 1.11 (0.79-1.56) |
| | Tapentadol SR | 87,542 | 70 | | 86,804 | 70 | |
| 28 days post-initiation | Oxycodone CR | 281,697 | 411 | 1.10 (0.90-1.33) | 86,804 | 128 | 1.09 (0.86-1.39) |
| | Tapentadol SR | 87,542 | 140 | | 86,804 | 140 | |

[1] Approximated by odds ratio

SR = sustained release; CR = controlled release; RR = relative risk; CI = confidence interval

**Supplementary Table 11. Baseline characteristics of tapentadol (SR) and oxycodone (CR) initiators with only study opioids dispensed on initiation date and no exposure to non-study opioids in the previous 90 days, before and after propensity score matching**

| Characteristic | Unmatched | | | Matched | | |
|---|---|---|---|---|---|---|
| | Tapentadol SR N = 56,947 | Oxycodone CR N = 171,359 | SD | Tapentadol SR N = 56,293 | Oxycodone CR N = 56,293 | SD |
| Year of initiation* | | | | | | |
| 2014 | 1,289 (2.3%) | 9,636 (5.6%) | .17 | 1,288 (2.3%) | 1,288 (2.3%) | .00 |
| 2015 | 4,695 (8.2%) | 30,861 (18.0%) | .29 | 4,690 (8.3%) | 4,690 (8.3%) | .00 |
| 2016 | 5,729 (10.1%) | 34,130 (19.9%) | .28 | 5,718 (10.2%) | 5,718 (10.2%) | .00 |
| 2017 | 9,446 (16.6%) | 34,389 (20.1%) | .09 | 9,438 (16.8%) | 9,438 (16.8%) | .00 |
| 2018 | 10,952 (19.2%) | 28,019 (16.4%) | .08 | 10,922 (19.4%) | 10,922 (19.4%) | .00 |
| 2019 | 14,060 (24.7%) | 21,652 (12.6%) | .31 | 13,959 (24.8%) | 13,959 (24.8%) | .00 |
| 2020 | 10,776 (18.9%) | 12,672 (7.4%) | .35 | 10,278 (18.3%) | 10,278 (18.3%) | .00 |
| Sex | | | | | | |
| Female | 33,443 (58.7%) | 95,072 (55.5%) | .07 | 32,995 (58.6%) | 33,244 (59.1%) | .01 |
| Male | 23,504 (41.3%) | 76,287 (44.5%) | | 23,298 (41.4%) | 23,049 (40.9%) | |
| Age group (years) | | | | | | |
| 18-44 | 10,780 (18.9%) | 33,623 (19.6%) | .02 | 10,717 (19.0%) | 11,256 (20.0%) | .02 |
| 45-64 | 19,549 (34.3%) | 51,926 (30.3%) | .09 | 19,241 (34.2%) | 18,947 (33.7%) | .01 |
| 65-84 | 23,226 (40.8%) | 68,187 (39.8%) | .02 | 22,959 (40.8%) | 22,400 (39.8%) | .02 |
| 85 plus | 3,392 (6.0%) | 17,623 (10.3%) | .16 | 3,376 (6.0%) | 3,690 (6.6%) | .02 |
| Remoteness Area | | | | | | |
| Major cities | 38,552 (67.7%) | 114,502 (66.8%) | .02 | 38,137 (67.8%) | 38,101 (67.7%) | .00 |
| Inner regional | 14,357 (25.2%) | 41,855 (24.4%) | .02 | 14,143 (25.1%) | 14,070 (25.0%) | .00 |
| Outer regional | 3,543 (6.2%) | 13,047 (7.6%) | .05 | 3,521 (6.3%) | 3,630 (6.5%) | .01 |
| Remote | 213 (0.4%) | 1,033 (0.6%) | .03 | 212 (0.4%) | 229 (0.4%) | .00 |
| Very remote | 31 (0.1%) | 97 (0.1%) | .00 | 30 (0.1%) | 25 (0.0%) | .00 |
| Missing | 251 (0.4%) | 825 (0.5%) | .01 | 250 (0.4%) | 238 (0.4%) | .00 |
| Decile of relative disadvantage | | | | | | |
| 1 (most disadvantage) | 4,008 (7.0%) | 12,936 (7.6%) | .02 | 3,979 (7.1%) | 3,831 (6.8%) | .01 |

| Characteristic | Unmatched | | | Matched | | |
|---|---|---|---|---|---|---|
| | Tapentadol SR N = 56,947 | Oxycodone CR N = 171,359 | SD | Tapentadol SR N = 56,293 | Oxycodone CR N = 56,293 | SD |
| 2 | 4,883 (8.6%) | 14,841 (8.7%) | .00 | 4,823 (8.6%) | 4,862 (8.6%) | .00 |
| 3 | 4,352 (7.6%) | 13,746 (8.0%) | .01 | 4,315 (7.7%) | 4,270 (7.6%) | .00 |
| 4 | 6,360 (11.2%) | 20,596 (12.0%) | .03 | 6,295 (11.2%) | 6,521 (11.6%) | .01 |
| 5 | 7,430 (13.1%) | 18,632 (10.9%) | .07 | 7,268 (12.9%) | 7,108 (12.6%) | .01 |
| 6 | 6,461 (11.4%) | 19,091 (11.1%) | .01 | 6,394 (11.4%) | 6,331 (11.3%) | .00 |
| 7 | 5,842 (10.3%) | 17,534 (10.2%) | .00 | 5,771 (10.3%) | 5,838 (10.4%) | .00 |
| 8 | 5,080 (8.9%) | 15,714 (9.2%) | .01 | 5,036 (9.0%) | 5,137 (9.1%) | .01 |
| 9 | 6,785 (11.9%) | 20,357 (11.9%) | .00 | 6,719 (11.9%) | 6,743 (12.0%) | .00 |
| 10 (least disadvantage) | 5,482 (9.6%) | 17,059 (10.0%) | .01 | 5,431 (9.7%) | 5,407 (9.6%) | .00 |
| Missing | 264 (0.5%) | 853 (0.5%) | .00 | 262 (0.5%) | 245 (0.4%) | .00 |
| Comorbidities | | | | | | |
| Cancer | 4,939 (8.7%) | 21,643 (12.6%) | .13 | 4,917 (8.7%) | 4,592 (8.2%) | .02 |
| Substance use disorder | 1,912 (3.4%) | 6,018 (3.5%) | .01 | 1,895 (3.4%) | 1,935 (3.4%) | .00 |
| Prior overdose | 85 (0.2%) | 253 (0.2%) | .00 | 83 (0.2%) | 79 (0.1%) | .00 |
| Depression | 16,902 (29.7%) | 45,941 (26.8%) | .06 | 16,608 (29.5%) | 16,918 (30.1%) | .01 |
| Anxiety | 5,716 (10.0%) | 15,980 (9.3%) | .02 | 5,632 (10.0%) | 5,408 (9.6%) | .01 |
| Hypertension | 11,666 (20.5%) | 35,701 (20.8%) | .01 | 11,466 (20.4%) | 11,990 (21.3%) | .02 |
| Atrial fibrillation or flutter | 1,206 (2.1%) | 5,339 (3.1%) | .06 | 1,199 (2.1%) | 1,234 (2.2%) | .00 |
| Diabetes | 8,575 (15.1%) | 27,914 (16.3%) | .03 | 8,474 (15.1%) | 8,704 (15.5%) | .01 |
| Congestive heart failure | 7,538 (13.2%) | 27,269 (15.9%) | .08 | 7,458 (13.3%) | 7,792 (13.8%) | .02 |
| Thyroid disease | 4,874 (8.6%) | 14,624 (8.5%) | .00 | 4,799 (8.5%) | 4,969 (8.8%) | .01 |
| Postural hypotension | 290 (0.5%) | 1,117 (0.7%) | .02 | 286 (0.5%) | 301 (0.5%) | .00 |
| Osteoporosis | 7,067 (12.4%) | 25,850 (15.1%) | .08 | 7,006 (12.5%) | 6,941 (12.3%) | .00 |
| Stroke | 11,852 (20.8%) | 40,530 (23.7%) | .07 | 11,744 (20.9%) | 11,115 (19.7%) | .03 |
| Renal impairment | 832 (1.5%) | 4,198 (2.5%) | .07 | 823 (1.5%) | 803 (1.4%) | .00 |
| Hepatic impairment | 413 (0.7%) | 2,214 (1.3%) | .06 | 411 (0.7%) | 318 (0.6%) | .02 |
| Surgery (prior 30 days) | 11,295 (19.8%) | 36,580 (21.4%) | .04 | 11,149 (19.8%) | 10,378 (18.4%) | .03 |

| Characteristic | Unmatched | | | Matched | | |
| --- | --- | --- | --- | --- | --- | --- |
| | Tapentadol SR N = 56,947 | Oxycodone CR N = 171,359 | SD | Tapentadol SR N = 56,293 | Oxycodone CR N = 56,293 | SD |
| Falls-risk medicines (FRIDs; prior 30 days) | | | | | | |
| Yes | 26,981 (47.4%) | 81,561 (47.6%) | .00 | 26,604 (47.3%) | 26,788 (47.6%) | .01 |
| Number of FRIDs: | | | | | | |
| 1 | 16,129 (59.8%) | 46,624 (57.2%) | .05 | 15,911 (59.8%) | 15,459 (57.7%) | .04 |
| 2 | 7,410 (27.5%) | 23,035 (28.2%) | .02 | 7,304 (27.5%) | 7,575 (28.3%) | .02 |
| 3 | 2,594 (9.6%) | 8,710 (10.7%) | .04 | 2,547 (9.6%) | 2,761 (10.3%) | .02 |
| 4 | 666 (2.5%) | 2,503 (3.1%) | .04 | 660 (2.5%) | 782 (2.9%) | .03 |
| 5+ | 182 (0.7%) | 689 (0.8%) | .02 | 182 (0.7%) | 211 (0.8%) | .01 |
| Type of FRID | | | | | | |
| *Medicines associated with high risk of falls* | | | | | | |
| Antipsychotics (excluding lithium) | 729 (2.7%) | 2,956 (3.6%) | .05 | 718 (2.7%) | 962 (3.6%) | .05 |
| Anxiolytics | 1,334 (4.9%) | 4,010 (4.9%) | .00 | 1,314 (4.9%) | 1,374 (5.1%) | .01 |
| Hypnotics and sedatives | 1,084 (4.0%) | 3,745 (4.6%) | .03 | 1,072 (4.0%) | 1,037 (3.9%) | .01 |
| Antidepressants | 9,118 (33.8%) | 25,832 (31.7%) | .05 | 8,945 (33.6%) | 9,681 (36.1%) | .05 |
| *Medicines that cause orthostatism/hypotension* | | | | | | |
| Vasodilators used in cardiac disease (e.g. nitrates) | 745 (2.8%) | 3,312 (4.1%) | .07 | 740 (2.8%) | 854 (3.2%) | .02 |
| Antihypertensives | 1,102 (4.1%) | 3,143 (3.9%) | .01 | 1,086 (4.1%) | 1,044 (3.9%) | .01 |
| Diuretics | 1,701 (6.3%) | 6,979 (8.6%) | .09 | 1,688 (6.3%) | 1,917 (7.2%) | .03 |
| Beta blockers | 4,178 (15.5%) | 14,311 (17.6%) | .06 | 4,141 (15.6%) | 4,310 (16.1%) | .01 |
| Calcium channel blockers | 4,495 (16.7%) | 14,704 (18.0%) | .04 | 4,429 (16.7%) | 4,558 (17.0%) | .01 |
| Renin-angiotensin acting agents | 16,621 (61.6%) | 49,576 (60.8%) | .02 | 16,393 (61.6%) | 16,165 (60.3%) | .03 |
| Alpha adrenoreceptor blockers | 631 (2.3%) | 2,018 (2.5%) | .01 | 623 (2.3%) | 588 (2.2%) | .01 |
| Dopaminergic agents | 598 (2.2%) | 1,818 (2.2%) | .00 | 592 (2.2%) | 626 (2.3%) | .01 |
| Other medicines associated with falls (prior 30 days) | | | | | | |
| Gabapentinoids | 5,082 (8.9%) | 9,672 (5.6%) | .13 | 4,861 (8.6%) | 4,575 (8.1%) | .02 |
| Anticholinergic medicines | 17 (0.0%) | 75 (0.0%) | .01 | 17 (0.0%) | 11 (0.0%) | .01 |
| Statins | 14,638 (25.7%) | 43,867 (25.6%) | .00 | 14,419 (25.6%) | 14,505 (25.8%) | .00 |

| Characteristic | Unmatched | | | Matched | | |
|---|---|---|---|---|---|---|
| | Tapentadol SR N = 56,947 | Oxycodone CR N = 171,359 | SD | Tapentadol SR N = 56,293 | Oxycodone CR N = 56,293 | SD |
| Hospital frailty risk score | | | | | | |
| Low: <5 | 51,001 (89.6%) | 145,753 (85.1%) | .14 | 50,387 (89.5%) | 50,374 (89.5%) | .00 |
| Medium: 5-15 | 4,527 (8.0%) | 18,085 (10.6%) | .09 | 4,499 (8.0%) | 4,403 (7.8%) | .01 |
| High: >15 | 1,419 (2.5%) | 7,521 (4.4%) | .10 | 1,407 (2.5%) | 1,516 (2.7%) | .01 |
| History of falls | | | | | | |
| Falls in the past 2 years to 60 days | 1,938 (3.4%) | 8,258 (4.8%) | .07 | 1,920 (3.4%) | 2,045 (3.6%) | .01 |

Note: cohort matched on propensity score, year and month of initiation

* study period: 01 September 2014 – 03 December 2020

SR = sustained release; CR = controlled release; SD = absolute standardised difference

**Supplementary Table 12. Relative risks of falls and cataract surgeries among tapentadol (SR) and oxycodone (CR) initiators with only study opioids dispensed on initiation date and no exposure to non-study opioids in the prior 90 days**

| Outcome | Exposure | Unmatched | | | Matched | | |
|---|---|---|---|---|---|---|---|
| | | No. exposed | No. events | RR (95% CI) | No. exposed | No. events | Matched RR[1] (95% CI) |
| Falls resulting in ED presentation, hospitalisation, or death | | | | | | | |
| 7 days post-initiation | Oxycodone CR | 171,359 | 547 | 0.59 (0.48-0.73) | 56,293 | 165 | 0.64 (0.50-0.81) |
| | Tapentadol SR | 56,947 | 108 | | 56,293 | 105 | |
| 14 days post-initiation | Oxycodone CR | 171,359 | 846 | 0.56 (0.47-0.67) | 56,293 | 247 | 0.63 (0.51-0.77) |
| | Tapentadol SR | 56,947 | 158 | | 56,293 | 155 | |
| 28 days post-initiation | Oxycodone CR | 171,359 | 1,213 | 0.60 (0.52-0.68) | 56,293 | 347 | 0.68 (0.57-0.80) |
| | Tapentadol SR | 56,947 | 240 | | 56,293 | 235 | |
| Cataract surgeries (negative control outcome) | | | | | | | |
| 7 days post-initiation | Oxycodone CR | 171,359 | 57 | 0.90 (0.52-1.54) | 56,293 | 14 | 1.21 (0.60-2.46) |
| | Tapentadol SR | 56,947 | 17 | | 56,293 | 17 | |
| 14 days post-initiation | Oxycodone CR | 171,359 | 122 | 0.89 (0.61-1.29) | 56,293 | 40 | 0.90 (0.57-1.41) |
| | Tapentadol SR | 56,947 | 36 | | 56,293 | 36 | |
| 28 days post-initiation | Oxycodone CR | 171,359 | 246 | 1.02 (0.79-1.30) | 56,293 | 79 | 1.05 (0.77-1.43) |
| | Tapentadol SR | 56,947 | 83 | | 56,293 | 83 | |

[1] Approximated by odds ratio

SR = sustained release; CR = controlled release; RR = relative risk; CI = confidence interval

**Supplementary Table 13. Baseline characteristics of tapentadol (SR) and oxycodone (CR) initiators between 1 September 2014 - 3 December 2020 who concurrently initiated oxycodone (IR), before and after propensity score matching**

| Characteristic | Unmatched | | | Matched | | |
|---|---|---|---|---|---|---|
| | Tapentadol SR N = 8,470 | Oxycodone CR N = 88,239 | SD | Tapentadol SR N = 8,436 | Oxycodone CR N = 8,436 | SD |
| Year of initiation* | | | | | | |
| 2014 | 36 (0.4%) | 5,298 (6.0%) | .32 | 35 (0.4%) | 35 (0.4%) | .00 |
| 2015 | 207 (2.4%) | 15,897 (18.0%) | .53 | 206 (2.4%) | 206 (2.4%) | .00 |
| 2016 | 525 (6.2%) | 17,316 (19.6%) | .41 | 523 (6.2%) | 523 (6.2%) | .00 |
| 2017 | 1,393 (16.5%) | 17,839 (20.2%) | .10 | 1,390 (16.5%) | 1,390 (16.5%) | .00 |
| 2018 | 2,156 (25.5%) | 14,490 (16.4%) | .22 | 2,150 (25.5%) | 2,150 (25.5%) | .00 |
| 2019 | 2,745 (32.4%) | 11,638 (13.2%) | .47 | 2,739 (32.5%) | 2,739 (32.5%) | .00 |
| 2020 | 1,408 (16.6%) | 5,761 (6.5%) | .32 | 1,393 (16.5%) | 1,393 (16.5%) | .00 |
| Sex | | | | | | |
| Female | 4,090 (48.3%) | 43,011 (48.7%) | .01 | 4,074 (48.3%) | 4,048 (48.0%) | .01 |
| Male | 4,380 (51.7%) | 45,228 (51.3%) | | 4,362 (51.7%) | 4,388 (52.0%) | |
| Age group (years) | | | | | | |
| 18-44 | 2,095 (24.7%) | 20,224 (22.9%) | .04 | 2,091 (24.8%) | 2,177 (25.8%) | .02 |
| 45-64 | 3,080 (36.4%) | 30,954 (35.1%) | .03 | 3,071 (36.4%) | 3,020 (35.8%) | .01 |
| 65-84 | 3,135 (37.0%) | 33,468 (37.9%) | .02 | 3,126 (37.1%) | 3,090 (36.6%) | .01 |
| 85 plus | 160 (1.9%) | 3,593 (4.1%) | .13 | 148 (1.8%) | 149 (1.8%) | .00 |
| Remoteness Area | | | | | | |
| Major cities | 5,591 (66.0%) | 58,115 (65.9%) | .00 | 5,571 (66.0%) | 5,651 (67.0%) | .02 |
| Inner regional | 2,286 (27.0%) | 23,084 (26.2%) | .02 | 2,275 (27.0%) | 2,232 (26.5%) | .01 |
| Outer regional | 503 (5.9%) | 6,239 (7.1%) | .05 | 501 (5.9%) | 485 (5.8%) | .01 |
| Remote | 40-45 | 345 (0.4%) | .01 | 39-44 | 30 (0.4%) | .02 |
| Very remote | <6 | 41 (0.1%) | ** | <6 | 6 (0.1%) | ** |
| Missing | 45 (0.5%) | 415 (0.5%) | .01 | 45 (0.5%) | 32 (0.4%) | .02 |
| Decile of relative disadvantage | | | | | | |
| 1 (most disadvantage) | 420 (5.0%) | 5,187 (5.9%) | .04 | 418 (5.0%) | 424 (5.0%) | .00 |

| Characteristic | Unmatched | | | Matched | | |
|---|---|---|---|---|---|---|
| | Tapentadol SR N = 8,470 | Oxycodone CR N = 88,239 | SD | Tapentadol SR N = 8,436 | Oxycodone CR N = 8,436 | SD |
| 2 | 591 (7.0%) | 7,302 (8.3%) | .05 | 591 (7.0%) | 582 (6.9%) | .00 |
| 3 | 542 (6.4%) | 6,838 (7.8%) | .05 | 540 (6.4%) | 559 (6.6%) | .01 |
| 4 | 891 (10.5%) | 10,431 (11.8%) | .04 | 888 (10.5%) | 923 (10.9%) | .01 |
| 5 | 898 (10.6%) | 9,361 (10.6%) | .00 | 895 (10.6%) | 812 (9.6%) | .03 |
| 6 | 926 (10.9%) | 10,282 (11.7%) | .02 | 923 (10.9%) | 945 (11.2%) | .01 |
| 7 | 879 (10.4%) | 8,807 (10.0%) | .01 | 874 (10.4%) | 850 (10.1%) | .01 |
| 8 | 902 (10.7%) | 9,071 (10.3%) | .01 | 900 (10.7%) | 940 (11.1%) | .02 |
| 9 | 1,351 (16.0%) | 10,877 (12.3%) | .10 | 1,340 (15.9%) | 1,287 (15.3%) | .02 |
| 10 (least disadvantage) | 1,021 (12.1%) | 9,657 (10.9%) | .03 | 1,018 (12.1%) | 1,079 (12.8%) | .02 |
| Missing | 49 (0.6%) | 426 (0.5%) | .01 | 49 (0.6%) | 35 (0.4%) | .02 |
| Exposure to non-study opioids (prior 90 days) | | | | | | |
| None | 7,085 (83.7%) | 70,507 (79.9%) | .10 | 7,071 (83.8%) | 7,071 (83.8%) | .00 |
| Yes | 1,385 (16.4%) | 17,732 (20.1%) | | 1,365 (16.2%) | 1,365 (16.2%) | |
| Comorbidities | | | | | | |
| Cancer | 678 (8.0%) | 11,954 (13.6%) | .18 | 666 (7.9%) | 675 (8.0%) | .00 |
| Substance use disorder | 302 (3.6%) | 3,481 (3.9%) | .02 | 301 (3.6%) | 301 (3.6%) | .00 |
| Prior overdose | <6 | 146 (0.2%) | ** | <6 | <6 | ** |
| Depression | 1,904 (22.5%) | 22,296 (25.3%) | .07 | 1,894 (22.5%) | 1,933 (22.9%) | .01 |
| Anxiety | 726 (8.6%) | 8,179 (9.3%) | .02 | 722 (8.6%) | 695 (8.2%) | .01 |
| Hypertension | 1,863 (22.0%) | 19,503 (22.1%) | .00 | 1,842 (21.8%) | 1,953 (23.2%) | .03 |
| Atrial fibrillation or flutter | 151 (1.8%) | 2,158 (2.5%) | .05 | 148 (1.8%) | 159 (1.9%) | .01 |
| Diabetes | 1,008 (11.9%) | 12,447 (14.1%) | .07 | 1,002 (11.9%) | 1,022 (12.1%) | .01 |
| Congestive heart failure | 750 (8.9%) | 10,209 (11.6%) | .09 | 739 (8.8%) | 771 (9.1%) | .01 |
| Thyroid disease | 541 (6.4%) | 6,240 (7.1%) | .03 | 539 (6.4%) | 554 (6.6%) | .01 |
| Postural hypotension | 31 (0.4%) | 422 (0.5%) | .02 | 30 (0.4%) | 30 (0.4%) | .00 |
| Osteoporosis | 801 (9.5%) | 11,034 (12.5%) | .10 | 791 (9.4%) | 819 (9.7%) | .01 |
| Stroke | 1,986 (23.5%) | 23,708 (26.9%) | .08 | 1,970 (23.4%) | 1,813 (21.5%) | .04 |
| Renal impairment | 58 (0.7%) | 1,424 (1.6%) | .09 | 54 (0.6%) | 52 (0.6%) | .00 |

| Characteristic | Unmatched | | | Matched | | |
|---|---|---|---|---|---|---|
| | Tapentadol SR N = 8,470 | Oxycodone CR N = 88,239 | SD | Tapentadol SR N = 8,436 | Oxycodone CR N = 8,436 | SD |
| Hepatic impairment | 60 (0.7%) | 1,096 (1.2%) | .05 | 58 (0.7%) | 45 (0.5%) | .02 |
| Surgery (prior 30 days) | 2,653 (31.3%) | 30,871 (35.0%) | .08 | 2,635 (31.2%) | 2,482 (29.4%) | .04 |
| Falls-risk medicines (FRIDs; prior 30 days) | | | | | | |
| Yes | 3,276 (38.7%) | 37,881 (42.9%) | .09 | 3,257 (38.6%) | 3,275 (38.8%) | .00 |
| Number of FRIDs: | | | | | | |
| 1 | 2,117 (64.6%) | 22,838 (60.3%) | .09 | 2,109 (64.8%) | 2,024 (61.8%) | .06 |
| 2 | 840 (25.6%) | 10,371 (27.4%) | .04 | 835 (25.6%) | 894 (27.3%) | .04 |
| 3 | 242 (7.4%) | 3,524 (9.3%) | .07 | 236 (7.3%) | 269 (8.2%) | .04 |
| 4 | 60 (1.8%) | 902 (2.4%) | .04 | 60 (1.8%) | 72 (2.2%) | .03 |
| 5+ | 17 (0.5%) | 246 (0.7%) | .02 | 17 (0.5%) | 16 (0.5%) | .00 |
| Type of FRID | | | | | | |
| *Medicines associated with high risk of falls* | | | | | | |
| Antipsychotics (excluding lithium) | 68 (2.1%) | 1,073 (2.8%) | .05 | 68 (2.1%) | 105 (3.2%) | .07 |
| Anxiolytics | 161 (4.9%) | 2,137 (5.6%) | .03 | 161 (4.9%) | 166 (5.1%) | .01 |
| Hypnotics and sedatives | 105 (3.2%) | 1,762 (4.7%) | .07 | 104 (3.2%) | 121 (3.7%) | .03 |
| Antidepressants | 1,042 (31.8%) | 12,672 (33.5%) | .04 | 1,036 (31.8%) | 1,052 (32.1%) | .01 |
| *Medicines that cause orthostatism/hypotension* | | | | | | |
| Vasodilators used in cardiac disease (e.g. nitrates) | 46 (1.4%) | 1,074 (2.8%) | .10 | 45 (1.4%) | 73 (2.2%) | .06 |
| Antihypertensives | 110 (3.4%) | 1,418 (3.7%) | .02 | 108 (3.3%) | 136 (4.2%) | .04 |
| Diuretics | 160 (4.9%) | 2,416 (6.4%) | .06 | 156 (4.8%) | 156 (4.8%) | .00 |
| Beta blockers | 431 (13.2%) | 5,655 (14.9%) | .05 | 427 (13.1%) | 465 (14.2%) | .03 |
| Calcium channel blockers | 514 (15.7%) | 6,396 (16.9%) | .03 | 510 (15.7%) | 519 (15.9%) | .01 |
| Renin-angiotensin acting agents | 2,090 (63.8%) | 23,001 (60.7%) | .06 | 2,079 (63.8%) | 2,042 (62.4%) | .03 |
| Alpha adrenoreceptor blockers | 67 (2.1%) | 812 (2.1%) | .01 | 65 (2.0%) | 96 (2.9%) | .06 |
| Dopaminergic agents | 57 (1.7%) | 620 (1.6%) | .01 | 56 (1.7%) | 58 (1.8%) | .00 |
| Other medicines associated with falls (prior 30 days) | | | | | | |
| Gabapentinoids | 626 (7.4%) | 5,571 (6.3%) | .04 | 615 (7.3%) | 564 (6.7%) | .02 |

| Characteristic | Unmatched | | | Matched | | |
|---|---|---|---|---|---|---|
| | Tapentadol SR N = 8,470 | Oxycodone CR N = 88,239 | SD | Tapentadol SR N = 8,436 | Oxycodone CR N = 8,436 | SD |
| Anticholinergic medicines | <6 | 23 (0.0%) | ** | <6 | <6 | ** |
| Statins | 1,811 (21.4%) | 19,632 (22.3%) | .02 | 1,797 (21.3%) | 1,770 (21.0%) | .01 |
| Hospital frailty risk score | | | | | | |
| Low: <5 | 7,739 (91.4%) | 76,838 (87.1%) | .14 | 7,718 (91.5%) | 7,732 (91.7%) | .01 |
| Medium: 5-15 | 583 (6.9%) | 8,852 (10.0%) | .11 | 573 (6.8%) | 547 (6.5%) | .01 |
| High: >15 | 148 (1.8%) | 2,549 (2.9%) | .08 | 145 (1.7%) | 157 (1.9%) | .01 |
| History of falls | | | | | | |
| Falls in the past 2 years to 60 days | 192 (2.3%) | 2,872 (3.3%) | .06 | 191 (2.3%) | 198 (2.4%) | .01 |

Note: cohort matched on propensity score, year and month of initiation, and exposure to non-study opioids in the prior 90 days

* study period: 01 September 2014 – 03 December 2020

** suppressed to avoid residual disclosure of small cells

SR = sustained release; CR = controlled release; IR = immediate release; SD = absolute standardised difference

**Supplementary Table 14. Relative risks of falls and cataract surgeries among tapentadol (SR) and oxycodone (CR) initiators who concurrently initiated oxycodone (IR)**

| Outcome | Exposure | Unmatched | | | Matched | | |
|---|---|---|---|---|---|---|---|
| | | No. exposed | No. events | RR (95% CI) | No. exposed | No. events | Matched RR[1] (95% CI) |
| Falls resulting in ED presentation, hospitalisation, or death | | | | | | | |
| 7 days post-initiation | Oxycodone CR | 88,239 | 257 | 0.57 (0.33-0.97) | 8,436 | 24 | 0.58 (0.30-1.13) |
| | Tapentadol SR | 8,470 | 14 | | 8,436 | 14 | |
| 14 days post-initiation | Oxycodone CR | 88,239 | 367 | 0.68 (0.45-1.03) | 8,436 | 30 | 0.80 (0.47-1.37) |
| | Tapentadol SR | 8,470 | 24 | | 8,436 | 24 | |
| 28 days post-initiation | Oxycodone CR | 88,239 | 498 | 0.65 (0.45-0.93) | 8,436 | 40 | 0.78 (0.49-1.24) |
| | Tapentadol SR | 8,470 | 31 | | 8,436 | 31 | |
| Cataract surgeries (negative control outcome) | | | | | | | |
| 7 days post-initiation | Oxycodone CR | 88,239 | 9 | 2.32 (0.50-10.71) | 8,436 | <6 | 1.00 (0.14-7.10) |
| | Tapentadol SR | 8,470 | <6 | | 8,436 | <6 | |
| 14 days post-initiation | Oxycodone CR | 88,239 | 24 | 1.30 (0.39-4.32) | 8,436 | <6 | 1.00 (0.20-4.96) |
| | Tapentadol SR | 8,470 | <6 | | 8,436 | <6 | |
| 28 days post-initiation | Oxycodone CR | 88,239 | 66 | 0.95 (0.41-2.18) | 8,436 | <6 | 1.20 (0.37-3.93) |
| | Tapentadol SR | 8,470 | 6 | | 8,436 | 6 | |

[1] Approximated by odds ratio

SR = sustained release; CR = controlled release; IR = immediate release; RR = relative risk; CI = confidence interval

**Supplementary Table 15. Average follow-up time (days), by exposure**

| Exposure | Unadjusted cohort | | | | | Matched cohort | | | | |
|---|---|---|---|---|---|---|---|---|---|---|
| | No. exposed | Mean | Std Dev | Min | Max | No. exposed | Mean | Std Dev | Min | Max |
| Oxycodone CR | 419,732 | 24.7 | 7.7 | 1 | 28 | 103,758 | 25.1 | 7.2 | 1 | 28 |
| Tapentadol SR | 103,924 | 25.3 | 7.1 | 1 | 28 | 103,758 | 25.3 | 7.1 | 1 | 28 |

Note: 99.5% censored

**Supplementary Table 16. Average follow-up time (days), by exposure, among initiators with no recent opioid exposure**

| Exposure | Unadjusted cohort | | | | | Matched cohort | | | | |
|---|---|---|---|---|---|---|---|---|---|---|
| | No. exposed | Mean | Std Dev | Min | Max | No. exposed | Mean | Std Dev | Min | Max |
| Oxycodone CR | 250,030 | 24.8 | 7.6 | 1 | 28 | 65,592 | 25.1 | 7.3 | 1 | 28 |
| Tapentadol SR | 65,715 | 25.0 | 7.3 | 1 | 28 | 65,592 | 25.0 | 7.3 | 1 | 28 |

Note: 99.5% censored

**Supplementary Table 17. Average follow-up time (days), by exposure, among initiators with recent exposure to opioids**

| Exposure | Unadjusted cohort | | | | | Matched cohort | | | | |
|---|---|---|---|---|---|---|---|---|---|---|
| | No. exposed | Mean | Std Dev | Min | Max | No. exposed | Mean | Std Dev | Min | Max |
| Oxycodone CR | 169,702 | 24.5 | 7.8 | 1 | 28 | 38,166 | 25.0 | 7.2 | 1 | 28 |
| Tapentadol SR | 38,209 | 25.6 | 6.6 | 1 | 28 | 38,166 | 25.6 | 6.6 | 1 | 28 |

Note: 99.4% censored

**Supplementary Table 18. E-values for relative risks of falls**

| Outcome | Exposure | No. exposed | No. events | Matched[1] RR | E-value | |
|---|---|---|---|---|---|---|
| | | | | | Estimate | CI[2] |
| Overall population | | | | | | |
| 7 days post-initiation | Oxycodone CR | 103,758 | 330 | 0.57 | 2.88 | 2.27 |
| | Tapentadol SR | 103,758 | 190 | | | |
| 14 days post-initiation | Oxycodone CR | 103,758 | 487 | 0.62 | 2.61 | 2.15 |
| | Tapentadol SR | 103,758 | 302 | | | |
| 28 days post-initiation | Oxycodone CR | 103,758 | 652 | 0.70 | 2.22 | 1.85 |
| | Tapentadol SR | 103,758 | 457 | | | |
| People aged 65 years and older | | | | | | |
| 7 days post-initiation | Oxycodone CR | 199,754 | 1,128 | 0.57 | 2.92 | 2.18 |
| | Tapentadol SR | 46,241 | 152 | | | |
| 14 days post-initiation | Oxycodone CR | 199,754 | 1,723 | 0.60 | 2.71 | 2.13 |
| | Tapentadol SR | 46,241 | 240 | | | |
| 28 days post-initiation | Oxycodone CR | 199,754 | 2,480 | 0.67 | 2.36 | 1.91 |
| | Tapentadol SR | 46,241 | 364 | | | |
| People aged 80 years and older | | | | | | |
| 7 days post-initiation | Oxycodone CR | 67,439 | 701 | 0.62 | 2.61 | 1.71 |
| | Tapentadol SR | 12,563 | 93 | | | |
| 14 days post-initiation | Oxycodone CR | 67,439 | 1,091 | 0.64 | 2.51 | 1.78 |
| | Tapentadol SR | 12,563 | 149 | | | |
| 28 days post-initiation | Oxycodone CR | 67,439 | 1,606 | 0.68 | 2.32 | 1.73 |
| | Tapentadol SR | 12,563 | 224 | | | |

[1] Approximated by odds ratio

[2] upper limit of the confidence interval presented as RR < 1

SR = sustained release; CR = controlled release; RR = relative risk; CI = confidence interval

**Supplementary Table 19. Relative risks of cataract surgeries (negative control outcome) for the total study population, people aged 65+, and people aged 80+**

| Outcome | Exposure | Unmatched | | | Matched | | | |
|---|---|---|---|---|---|---|---|---|
| | | No. exposed | No. events | RR (95% CI) | No. exposed | No. events | Matched RR[1] (95% CI) | Adjusted[2] RR[1] (95% CI) |
| Overall population | | | | | | | | |
| 7 days post-initiation | Oxycodone CR | 419,732 | 112 | 1.12 (0.75-1.66) | 103,758 | 22 | 1.41 (0.82-2.43) | 1.26 (0.69-2.31) |
| | Tapentadol SR | 103,924 | 31 | | 103,758 | 31 | | |
| 14 days post-initiation | Oxycodone CR | 419,732 | 242 | 1.25 (0.97-1.62) | 103,758 | 56 | 1.32 (0.93-1.87) | 1.16 (0.81-1.68) |
| | Tapentadol SR | 103,924 | 75 | | 103,758 | 74 | | |
| 28 days post-initiation | Oxycodone CR | 419,732 | 521 | 1.19 (1.00-1.43) | 103,758 | 117 | 1.31 (1.03-1.66) | 1.21 (0.95-1.56) |
| | Tapentadol SR | 103,924 | 154 | | 103,758 | 153 | | |
| People aged 65 years and older | | | | | | | | |
| 7 days post-initiation | Oxycodone CR | 199,754 | 93 | 1.11 (0.71-1.75) | 41,546 | 18 | 1.28 (0.69-2.37) | 1.06 (0.55-2.05) |
| | Tapentadol SR | 46,241 | 24 | | 41,546 | 23 | | |
| 14 days post-initiation | Oxycodone CR | 199,754 | 211 | 1.31 (0.99-1.73) | 41,546 | 52 | 1.10 (0.75-1.60) | 0.98 (0.66-1.46) |
| | Tapentadol SR | 46,241 | 64 | | 41,546 | 57 | | |
| 28 days post-initiation | Oxycodone CR | 199,754 | 450 | 1.27 (1.04-1.54) | 41,546 | 101 | 1.19 (0.91-1.55) | 1.13 (0.86-1.49) |
| | Tapentadol SR | 46,241 | 132 | | 41,546 | 120 | | |
| People aged 80 years and older | | | | | | | | |
| 7 days post-initiation | Oxycodone CR | 67,439 | 28 | 1.34 (0.59-3.07) | 10,783 | <6 | 1.75 (0.51-5.98) | 2.00 (0.50-8.00) |
| | Tapentadol SR | 12,563 | 7 | | 10,783 | 7 | | |
| 14 days post-initiation | Oxycodone CR | 67,439 | 67 | 1.44 (0.86-2.43) | 10,783 | 14 | 1.07 (0.52-2.22) | 1.07 (0.51-2.25) |
| | Tapentadol SR | 12,563 | 18 | | 10,783 | 15 | | |
| 28 days post-initiation | Oxycodone CR | 67,439 | 142 | 1.47 (1.04-2.10) | 10,783 | 26 | 1.23 (0.73-2.07) | 1.21 (0.72-2.04) |
| | Tapentadol SR | 12,563 | 39 | | 10,783 | 32 | | |

[1] Approximated by odds ratio

[2] Conditional logistic regression models adjusted for simultaneous initiation of study and non-study opioids

SR = sustained release; CR = controlled release; RR = relative risk; CI = confidence interval

**Supplementary Figure 1. Study design**

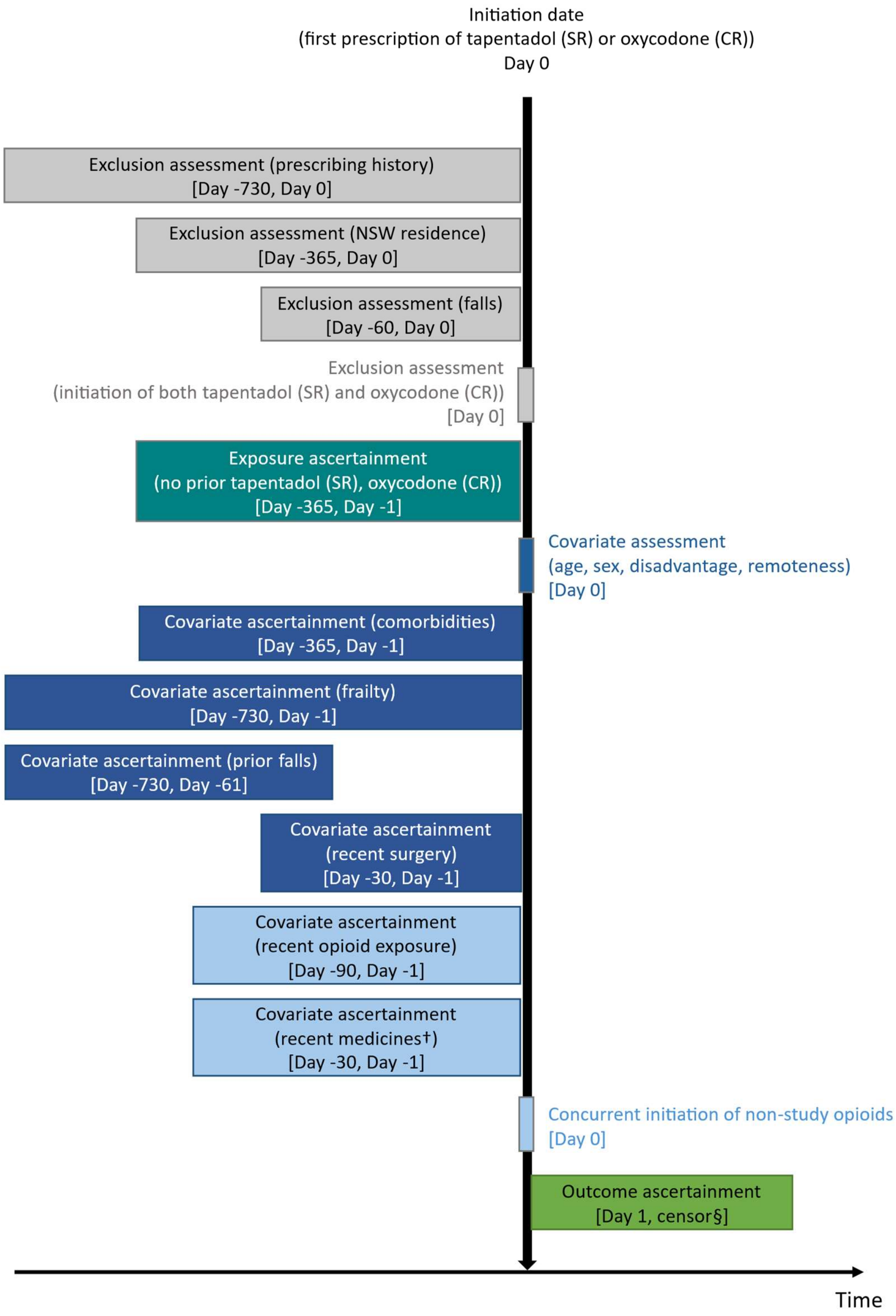


† Falls-risk medicines, gabapentinoids, anticholinergic medicines, statins

§ First of: outcome of interest, end of follow-up (7, 14, or 28 days), hospital admission (unrelated to falls), death, or study end date (31 December 2020)

**Supplementary Figure 2. Propensity score distributions for tapentadol (SR) and oxycodone (CR) initiators**

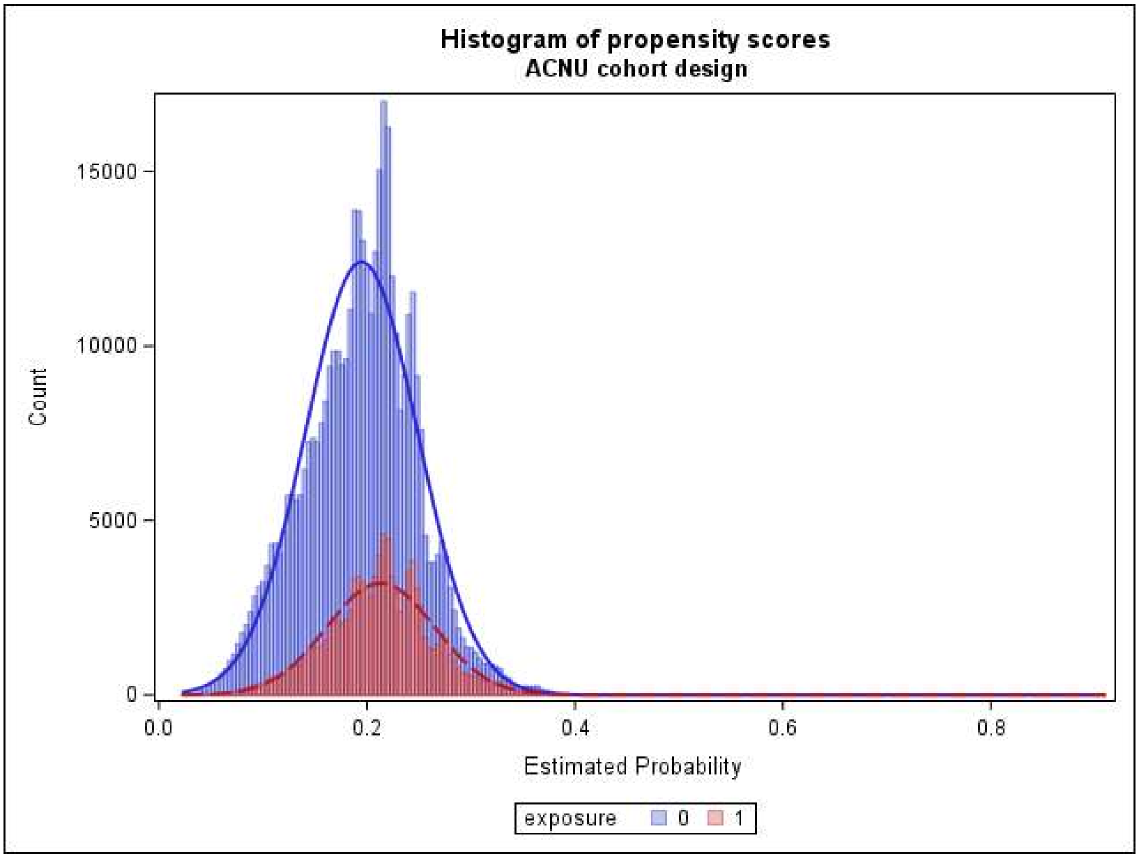

**Supplementary Figure 3. Cumulative incidence curves: falls following initiation of tapentadol (SR) or oxycodone (CR)**

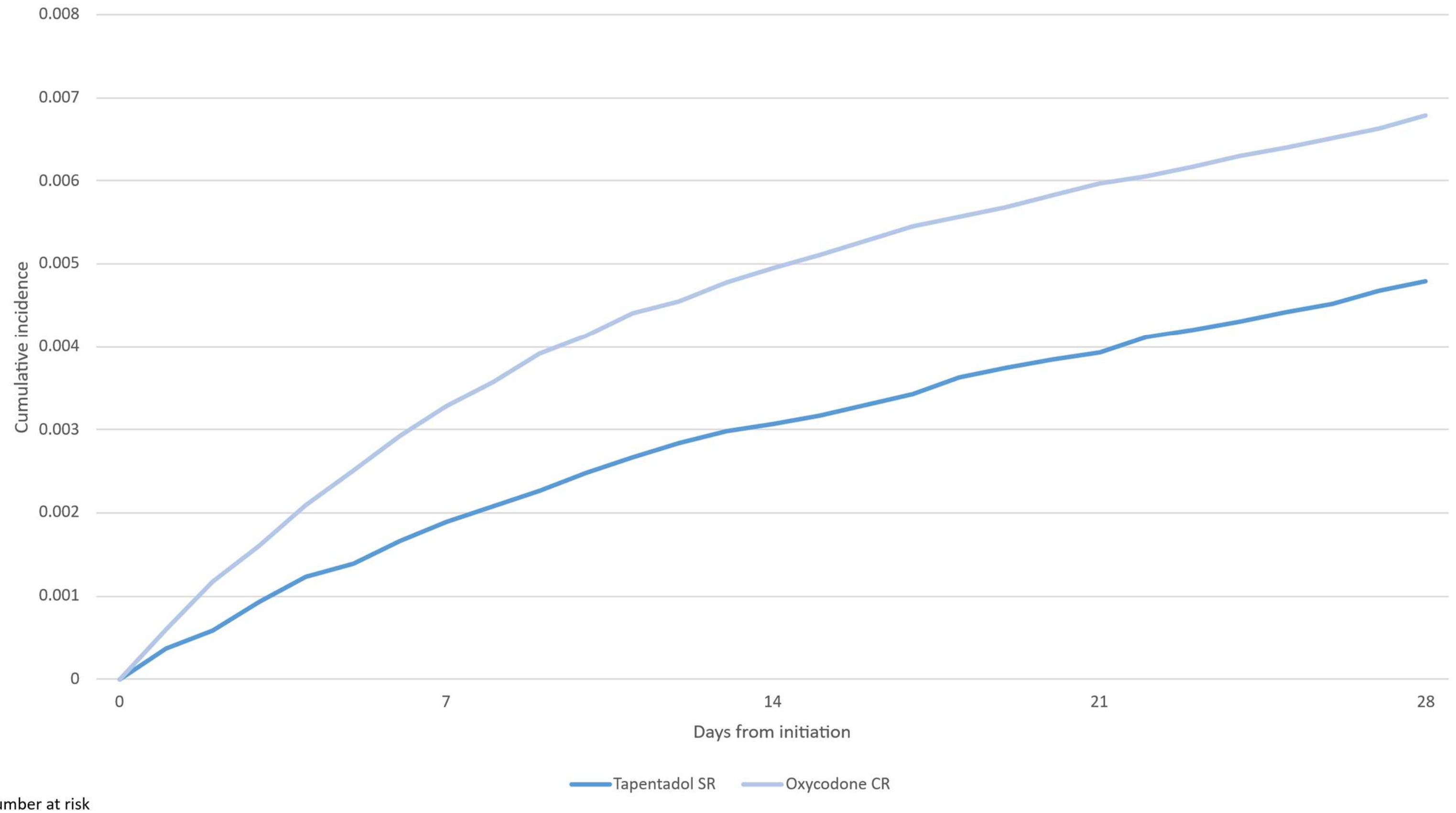


Number at risk

| | | | | | |
|---|---|---|---|---|---|
| Tapentadol | 103758 | 96900 | 92694 | 89965 | 87893 |
| Oxycodone | 103758 | 96603 | 92100 | 89264 | 87054 |

**Supplementary Figure 4. Cumulative incidence curves: falls following initiation of tapentadol (SR) or oxycodone (CR) among initiators with no recent opioid exposure**

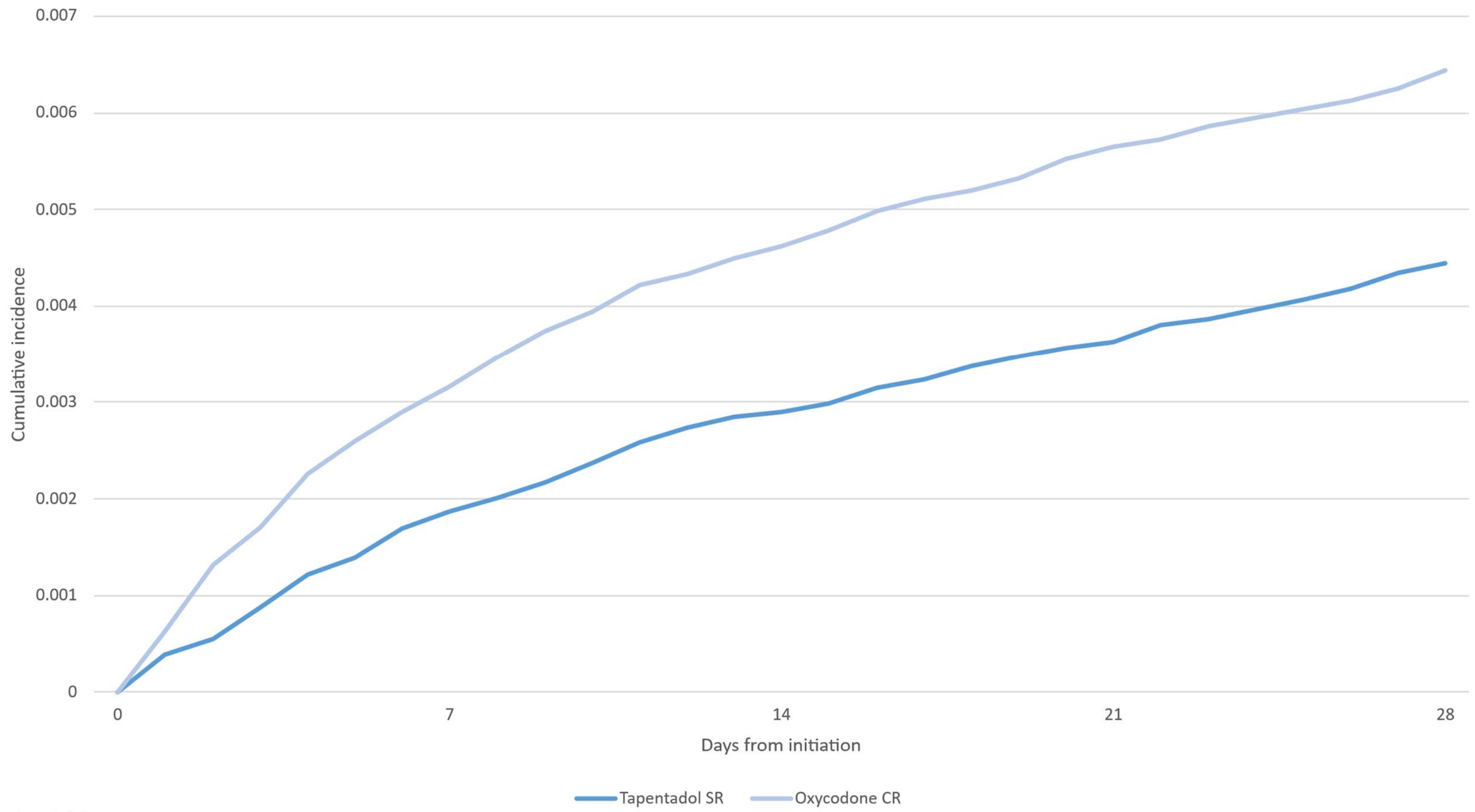


Number at risk

| | | | | | |
|---|---|---|---|---|---|
| Tapentadol | 65592 | 60767 | 57985 | 56262 | 55036 |
| Oxycodone | 65592 | 60966 | 58235 | 56584 | 55381 |

**Supplementary Figure 5. Cumulative incidence curves: falls following initiation of tapentadol (SR) or oxycodone (CR) among initiators with recent exposure to opioids**

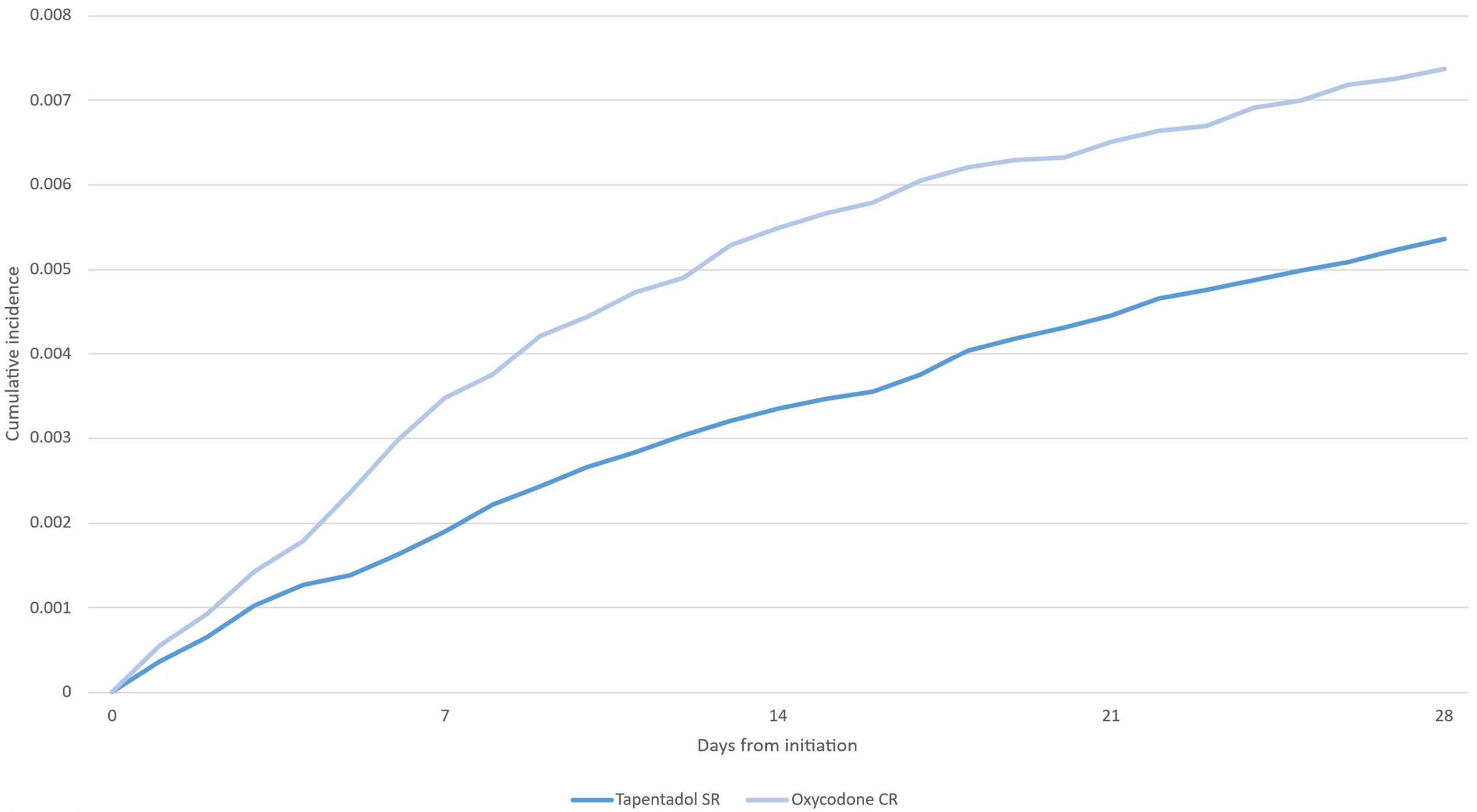


Number at risk

| | | | | | |
|---|---|---|---|---|---|
| Tapentadol | 38166 | 36133 | 34709 | 33703 | 32857 |
| Oxycodone | 38166 | 35637 | 33865 | 32680 | 31673 |
